# Biotypes of deeply phenotyped depressed patients reflect signatures of adverse childhood experience and depressive cognitive biases

**DOI:** 10.1101/2025.07.03.25330801

**Authors:** Federica Colombo, Federico Calesella, Beatrice Bravi, Lidia Fortaner-Uyà, Camilla Monopoli, Tommaso Cazzella, Michele Acconcia, Raffaella Zanardi, Cristina Colombo, Matteo Carminati, Chiara Fabbri, Alessandro Serretti, Irene Bollettini, Francesco Benedetti, Sara Poletti, Benedetta Vai

## Abstract

**Background:** Negative Cognitive Styles (NCS) are key features of depression contributing to severe clinical outcomes by sustaining negative affect. However, depression is clinically heterogeneous, reflecting complex neurobiological and environmental interactions. Characterizing heterogeneity using multimodal data could help identify mechanisms mapping onto different phenotypes and discover high-impact biomarkers.

**Methods:** Using a stability-based relative clustering validation pipeline, 344 depressed patients (135 major depressive disorder, 209 bipolar disorder) were stratified based on multimodal neuroimaging data, including grey matter volumes, cortical thickness, white matter diffusivity indices, resting-state functional connectivity (FC), and spontaneous neural activity. Clusters were derived from each modality separately and in combination, and profiled for Adverse Childhood Experiences (ACEs) and NCS.

**Results:** All neuroimaging modalities stratified depressed patients into two clusters, with the FC-based model achieving the highest accuracy (85%). Compared with healthy controls (HC, N=138), these clusters exhibited opposite patterns of global functional hyper-integration (Cluster 1) and hyper-segregation (Cluster 2). Unique multivariate neurobiological-ACEs relationships characterized the FC-based clusters. While ACEs negatively affected FC in Cluster 1 and HC, Cluster 2 showed opposite effects specific to ACE subtypes, with sexual abuse positively influencing FC. Mediation analysis showed that FC strength mediated the relationship between ACEs and NCS of overgeneralize only in Cluster 2. ACE-related functional alterations pinpoint brain regions involved in socioemotional and cognitive development, sensitive to maturational neural changes.

**Conclusions:** These findings provide evidence of clinically meaningful depression “biotypes” shaped by ACEs and associated with NCS, underlining the feasibility of computational psychiatry tools to uncover data-driven patterns for precision psychiatry.

## Introduction

Depressive disorders carry a substantial social and economic burden, demanding more personalized and effective clinical strategies (Cirone et al., 2021; GBD Mental Disorders Collaborators, 2022). Negative Cognitive Styles (NCS) - including dysfunctional attributional styles and negative thinking – are key prognostic factors in the onset and maintenance of depression (Beck, 2008). NCS are associated with poor therapeutic responses and increased suicidality (Fazakas-DeHoog et al., 2017; Shankman et al., 2013; Zhou et al., 2021), while targeted interventions can improve clinical symptoms and overall functioning (Jones and Sharpe, 2017; Scheepens et al., 2022; Spinhoven et al., 2018). Identifying the factors that influence and moderate NCS is of critical importance, particularly given the substantial heterogeneity in how patients may respond to treatments (Maslej et al., 2021).

It is widely recognised that environmental factors significantly contribute to the variance associated with mood disorders, and, by interacting with genetic predisposition, determine their clinical manifestation (Kendall et al., 2021). Adverse Childhood Experiences (ACEs), such as abuse and neglect, represents a key risk factor for psychopathology, including depression (McLaughlin et al., 2020; Poletti, Colombo, et al., 2014; Wade et al., 2022). Individuals exposed to ACEs often experience a worse illness course, including poor treatment response and increased suicidality (Blair et al., 2025; Giampetruzzi et al., 2023; Hayashi et al., 2015). Cognitive theories of depression propose that ACEs foster NCS by shaping early maladaptive cognitive patterns (Beck, 2008). This pathway could be potentially driven by ACEs-related structural and functional brain alterations (Teicher and Samson, 2013), which primarily impact networks governing emotion regulation, threat detection, and cognitive functions (Teicher et al., 2016; Yan et al., 2025). Exposure to ACEs was indeed shown to affect neural correlates specifically associated with psychiatric conditions, including grey matter (GM) volumetric reductions and white matter (WM) disruption (Benedetti et al., 2011, 2012, 2014; Poletti et al., 2014b, 2016, 2018), and associates with worse outcomes and higher suicidality (Vai, Serretti, et al., 2020).

Understanding how these factors moderate and predict clinical outcomes is a primary objective of precision psychiatry, where the identification of patient-specific characteristics enables the optimal prediction of intervention efficacy (Bzdok and Meyer-Lindenberg, 2018; Fernandes et al., 2017; Tiego et al., 2023). However, these interactions are unlikely to be consistent across patients, reflecting transdiagnostic heterogeneity (Xie et al., 2023). Recent research has identified neurobiological subtypes (i.e., biotypes) of depression, offering a data-driven stratification to better capture clinical and biological diversity. For instance, increased default mode network functional connectivity (FC) was linked to a “rumination” biotype (Goldstein-Piekarski et al., 2022; Williams, 2017, 2016), while heightened activity within the cognitive control circuit predicts negative biases and treatment response (Tozzi et al., 2024). Nevertheless, stratification studies have predominantly relied on resting-state functional MRI (Drysdale et al., 2017; Liang et al., 2020; Price et al., 2017; Tokuda et al., 2018), leaving the structural basis largely unexplored. We recently identified distinct biotypes of major depressive disorder (MDD) using structural neuroimaging: one marked by reduced GM, cortical thickness (CT), and WM integrity - linked to anergy, childhood abuse, and inflammatory markers - and another with more preserved structure, associated with cognitive symptoms (e.g., negative self-esteem) (Colombo et al., 2024). However, exploring interactions between ACEs and NCS in different biotypes for multimodal neuroimaging data remain unaddressed.

In this study we aimed to address these gaps by providing a unique neuroimaging-driven stratification through an unsupervised machine learning (ML) approach in depressed MDD and bipolar disorder (BD) patients, using multimodal structural and functional brain imaging data. We then examined the neurobiological and clinical features of each cluster, focusing on the relationships between NCS, ACEs, and neural correlates. Finally, we explored whether ACEs-related brain signatures mediates the relationship between ACEs and NCS using multivariate models, which are well-suited to capture complex brain-behavior associations underlying psychiatric heterogeneity (Sha et al., 2019; Xie et al., 2023).

## Methods and Materials

### Participants

The sample included 344 inpatients (135 MDD, 209 BD), consecutively admitted at the IRCCS San Raffaele Hospital (Milan, Italy) and diagnosed according DSM-5 criteria using the Structured Clinical Interview for the Diagnosis of Axis I Disorders (SCID-5). Inclusion criteria were age 18-65 years old and current depressive episode. Depression severity was assessed with the 21-items Hamilton Depression Rating Scale (HDRS-21) (Hamilton, 1986). Exclusion criteria for all patients included: major medical and neurological disorders, pregnancy, intellectual disability, history of drug or alcohol abuse or dependency, and psychiatric comorbidity on axis I. A sample of 138 healthy controls (HC) without psychiatric history or first-degree familial risk was also recruited. MRI acquisitions were performed for all participants between 2.30 pm and 6.00 pm. After a complete description of the study, written informed consent was obtained. All procedures contributing to this work comply with the ethical standards of the relevant national and institutional committees on human experimentation and with the Helsinki Declaration of 1975, as revised in 2008. The study was approved by the local ethical committee.

### Sociodemographic, childhood trauma and cognitive styles measures

Age, sex, number of previous mood episodes, age of onset, duration of illness, and years of education were collected. Medication load was calculated for each subject categorizing each medication into 0-4 scores based on dose range, and then combined to obtain a single composite score (see **Methods S1** for detailed criteria) (Sackeim, 2001). In a subsample of 229 patients (106 MDD, 123 BD) and 98 HC, childhood trauma was assessed through the 28-item version of the Childhood Trauma Questionnaire (CTQ) (Bernstein et al., 2003), which investigates 5 categories of childhood trauma: physical, emotional, and sexual abuse; physical and emotional neglect. Each form of maltreatment is evaluated through 5 items, where high scores reflect higher exposure (Bernstein et al., 2003). NCS were assessed through the Cognition Questionnaire (CQ) (Fennell and Campbell, 1984), which provides a fine-tuned evaluation of negative thinking in response to positive, neutral, and negative events (Fennell and Campbell, 1984). Five different dimensions of NCS can be derived: emotional impact, attribution of causality, generalization across time, generalization across situations, and perceived uncontrollability. High scores in each domain reflect high degree of that specific cognitive distortion.

### MRI data acquisition, processing and features extraction

The acquisition of T1-weighted, diffusion tensor imaging (DTI), and resting-state fMRI images was performed with two different 3.0 Tesla scanners. Details about acquisitions parameters for each scanner are reported in **Methods S2**.

The processing of T1-weighted neuroanatomical images was performed through the Computational Anatomy Toolbox (CAT12) for statistical parametric mapping (SPM) software (Gaser et al., 2024). The main pre-processing steps involved: normalization in the Montreal Neurological Institute (MNI) space; tissue segmentation into GM, WM, and cerebrospinal fluid (CSF); check of spatial alignment and sample homogeneity to exclude outliers; spatial smoothing of each GM map with a 6 mm full width at half maximum Gaussian filter. CT was computed for 68 regions derived from the Desikan-Killiany atlas (Desikan et al., 2006), while cortical and subcortical regional volumes were extracted for 122 regions identified from the Neuromorphometrics atlas (http://Neuromorphometrics.com/). The total intracranial volume (TIV) was also computed.

DTI pre-processing was performed using FMRIB Software Library (FSL) 6.0 tools (Smith et al., 2006, 2004; Woolrich et al., 2009). All images were corrected for eddy currents and head motion (Horsfield, 1999) and skull-stripped (Smith, 2002). Non-linear registration to the FMRIB58-FA template was performed, and a diffusion tensor model was fitted at each voxel to obtain voxel-wise maps of fractional anisotropy (FA), axial (AD), radial (RD), and mean diffusivity (MD). The obtained images were then processed using FSL’s Tract Based Spatial Statistics analytic method and aligned to the MNI space through the Non-Linear Image Registration Tool (FNIRT). For each DTI index, the maps of all subjects were merged into a common 4D image, and a “thinning” process was applied to create a skeletonized mean image representing the centers of all common tracts. A threshold of 0.2 was set for this image to control for inter-subject variability and reduce the likelihood of a partial volume effect. Whole-brain tract-based average values for each diffusion index were extracted according to ENIGMA-DTI protocols. Average FA values were calculated from voxels in each subject’s white matter skeleton within 43 tract-wise ROIs, derived from the Johns Hopkins University parcellation atlas (Mori et al., 2008). The same procedure was performed for the extraction of tract-based average values of AD, RD and MD. All measures were separately calculated for the right and left hemispheres in each tract, except for midline structures.

The T2*-weighted images were analyzed using the Harmonized AnaLysis of Functional MRI pipeline (HALFpipe) version 1.2.1 (Waller et al., 2022). Pre-processing steps encompass motion correction, slice time correction, susceptibility distortion correction, coregistration, and spatial normalization to the MNI152NLin2009cAsym template. Denoising was performed on the resampled images and included: spatial smoothing with a 60mm full-width at half-maximum kernel; grand mean scaling of 10,000; ICA-AROMA (Pruim et al., 2015a, 2015b); and temporal filtering using a Gaussian weighted high-pass (width of 125 s) for FC and a frequency-based bandpass (0.01-0.1 Hz) for measures of local neural activity. Physiological nuisance regressors were isolated for anatomical component correction (aCompCor) considering the top 5 CSF principal components (Muschelli et al., 2014). Several resting-state fMRI features were calculated. Considering FC, the time series from 246 Brainnetome (Fan et al., 2016) and 17 cerebellar regions of interest (ROIs) were extracted (Buckner et al., 2011). ROIs with missing time series across participants were excluded, resulting in 186 ROIs (**Table S1**). From the individual ROI-to-ROI FC matrices, positive (Spos) and negative (Sneg) strengths (i.e., the sum of positive and negative weights of links connected to a region) were computed for each ROI using the Python-equivalent of the Brain Connectivity Toolbox (bctpy, https://github.com/aestrivex/bctpy). Local activity metrics included the fractional amplitude of low frequency fluctuations (fALFF) and regional homogeneity (ReHo), calculated from the pre-processed images. fALFF was defined as the ratio of low frequencies (0.01–0.1 Hz) to the entire frequency power (Zou et al., 2008). ReHo measures the temporal synchronization of a voxel with its nearest neighboring voxels, calculated using the Kendall’s coefficient of concordance (Zang et al., 2004). Voxel-wise fALFF and ReHo maps for each participant were Z-standardized, and the corresponding average values across 186 ROIs were extracted.

To account for inter-scanner variability, ComBat harmonization (Johnson et al., 2007) was performed for each neuroimaging feature using neuroCombat (https://github.com/Jfortin1/ComBatHarmonization/tree/master), considering age, sex, diagnosis, and TIV (for GM and CT) as biological covariates to be preserved.

### Unsupervised machine learning analysis

To uncover biologically-driven subtypes of depression, a stability-based relative clustering validation algorithm (*NeuReval* Python package) (Colombo et al., 2024; Landi et al., 2021) was implemented for each neuroimaging modality (**Figure 1B**). This method leverages cross-validation to evaluate clustering generalizability by embedding unsupervised clustering in a supervised learning framework (Lange et al., n.d.). For all the models, input features were standardized, and confounding effects (age, sex, pharmacological load, frequency of episodes, and TIV for GM and CT) were regressed out, including age² and age-by-sex interaction to account for non-linear effects (Alfaro-Almagro et al., 2021). All these steps were fitted on the training set and then applied to the test set. A 2-fold cross-validation scheme (50% training, 50% test) was employed to account for size imbalances resulting from training-test sets splitting (Landi et al., 2021), and it was repeated 100 times to ensure robustness. For all the models, K-Means was implemented for clustering, while Support Vector Machine (SVM) served as supervised classifier. A grid search procedure was performed for hyperparameters tuning, using the minimization of the normalized stability as the criterion to select the optimal ones. These included K-Means initialization (random or k-means++), the soft-margin C (range: 0.01, 0.1, 1, 10, 100, 1000), and kernel type (linear or radial basis function) for SVM. Clustering solutions were evaluated for 2 to 5 clusters, and the one achieving the minimum normalized stability throughout cross-validation was selected.

To investigate whether the combination of different neuroimaging modalities improved stratification over single-modality models, a multiview spectral clustering algorithm was embedded within the *NeuReval* pipeline (https://github.com/fede-colombo/Multiview_NeuReval). In multiview frameworks, different feature sets (“views”) are entered within the clustering algorithm, which can be separately used to derive clusters. The rationale behind this approach is that the grouping structure identified in one view should agree with the one derived from another view. The best clustering solution is the one that assigns an observation to the same cluster regardless of the specific view (Kumar and Daumé, 2011). Only modalities that resulted in stable solutions in single-modality models were included, considering the same cross-validation scheme and confounds controls. Tuned hyperparameters included the affinity matrix (radial basis function or nearest neighbors) for multiview spectral clustering and SVM hyperparameters.

The internal quality of clustering solutions was further assessed with the Silhouette score and the Davies-Bouldin index. Statistical significance was tested with 10,000 Monte Carlo simulations using the *sigclust* method (R library) (Liu et al., 2008). Only stratification models that passed false-discovery rate (FDR) q<0.05 threshold were deemed as significant.

### Univariate and multivariate characterization of the clusters

For each significant model, clusters were compared for age, diagnosis, sex, number of previous episodes, years of education, age of onset, duration of illness, pharmacological load, HDRS-21 total score, CTQ and CQ total scores and domains. The same approach was employed for neuroimaging features. Cohen’s d effects sizes were calculated and multiple comparisons were FDR-corrected (q<0.05). Comparisons with HC were conducted only for the best-performing model. Cluster assignment agreement across models was assessed with the Fleiss’ kappa statistic.

For each significant stratification model, grouped partial least squares correlation (PLSC) analyses were also performed to investigate whether the multivariate neurobiological-ACEs relationship was different between clusters (myPLS MATLAB toolbox, https://github.com/danizoeller/myPLS/). (**Figure 1 D-E**). PLSC is a multivariate technique that identifies latent variable (LV) pairs capturing shared variance between two datasets (e.g., brain and behavioral data). With group labels, PLSC computes separate covariance matrices, enabling detection of between-cluster differences in multivariate brain–behavior associations (Krishnan et al., 2011). Prior to analyses, neuroimaging variables were residualized for the same confounds considered for *NeuReval*. CTQ scores were adjusted for age, sex, and education. For each model, 5000 permutations were performed to assess the statistical significance of the LV pairs (FDR q<0.05). Brain bootstrap ratio (BSR) and 95% confidence intervals (CI) on neurobiological-ACEs correlations were estimated through 5000 bootstrap resampling. BSRs were computed by dividing the correlation coefficient for each biological variable by its bootstrapped standard deviation, and interpreted as standardized weights (Krishnan et al., 2011). BSRs>|2| were judged as important in guiding the neurobiological-ACEs relationships, whereas CTQ loadings with 95% CI that do not include zero were deemed as significant. BSRs were converted into p-values and FDR-corrected (q<0.05) (Kebets et al., 2019).

For significant LV pairs, multivariate general linear models (GLMs) were implemented separately for each cluster, with LV neurobiological scores predicting CQ domains. Univariate analyses were also performed within each cluster to explore the effect of LV neurobiological scores on each CQ domain. Statistical significance was assessed using parametric estimates of the predictor variables, calculated by the least squares method. For those CQ domains that were significantly predicted by LV neurobiological scores, mediation models were performed, considering LV CTQ scores as independent variables, LV neurobiological scores as mediator, and CQ domain as dependent variable (5000 bootstraps, 90% CI) (SPSS PROCESS macro) (Hayes, 2012). (**Figure 1F**).

## Results

Descriptive statistics of the whole sample (N=482) are reported in **Table 1**. No significant differences in age, sex, duration of illness, and pharmacological load were observed between MDD and BD. However, BD patients showed a significantly higher number of previous episodes compared to MDD (q<0.001). BD and MDD patients were also comparable for HDRS-21, CQ and CTQ domains. HC were younger, with higher level of education and lower CTQ and CQ scores compared to both MDD and BD. In the whole sample, significant and positive correlations were found across all the CTQ and CQ domains (**Figure S1**).

**Table 1.**
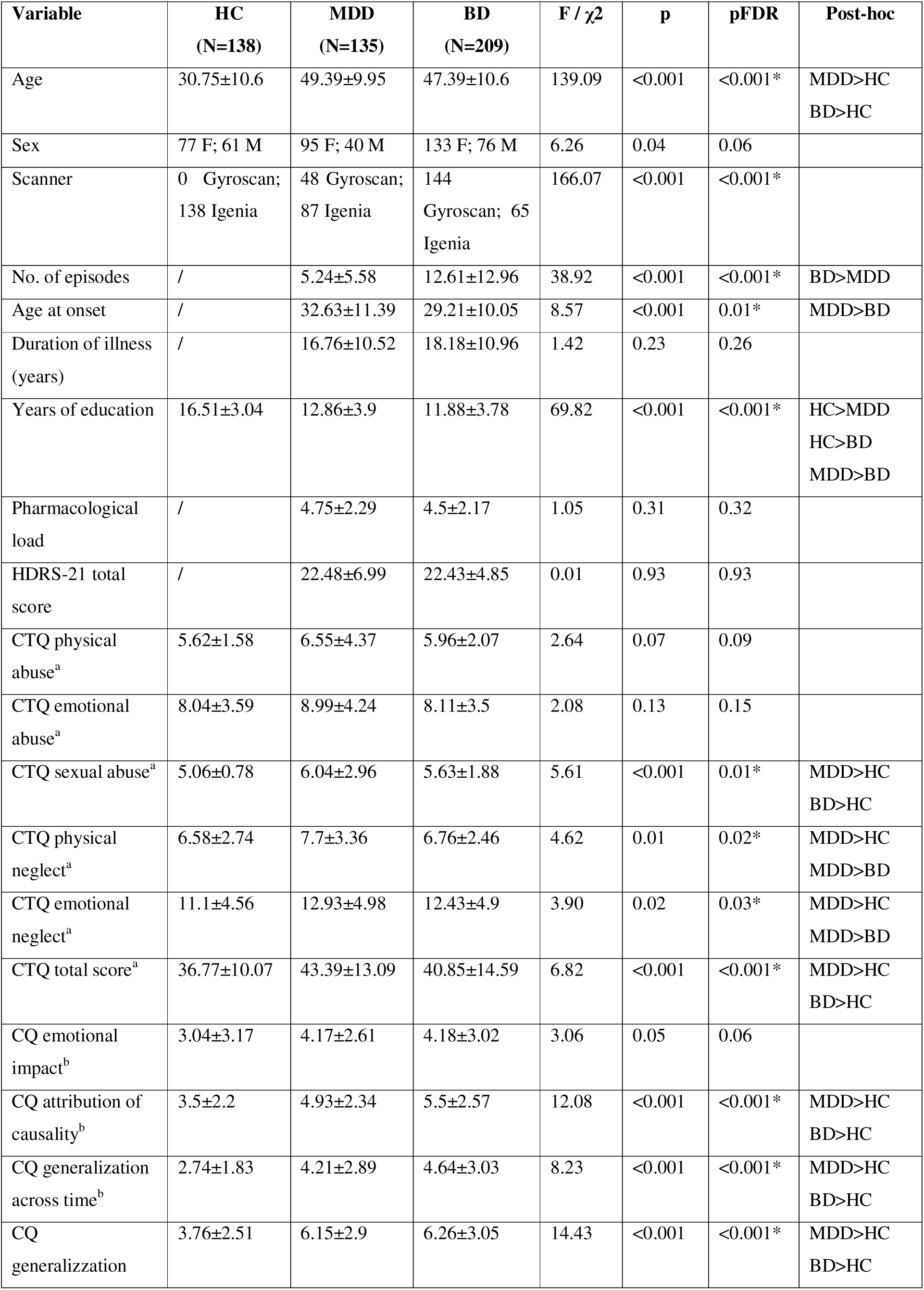
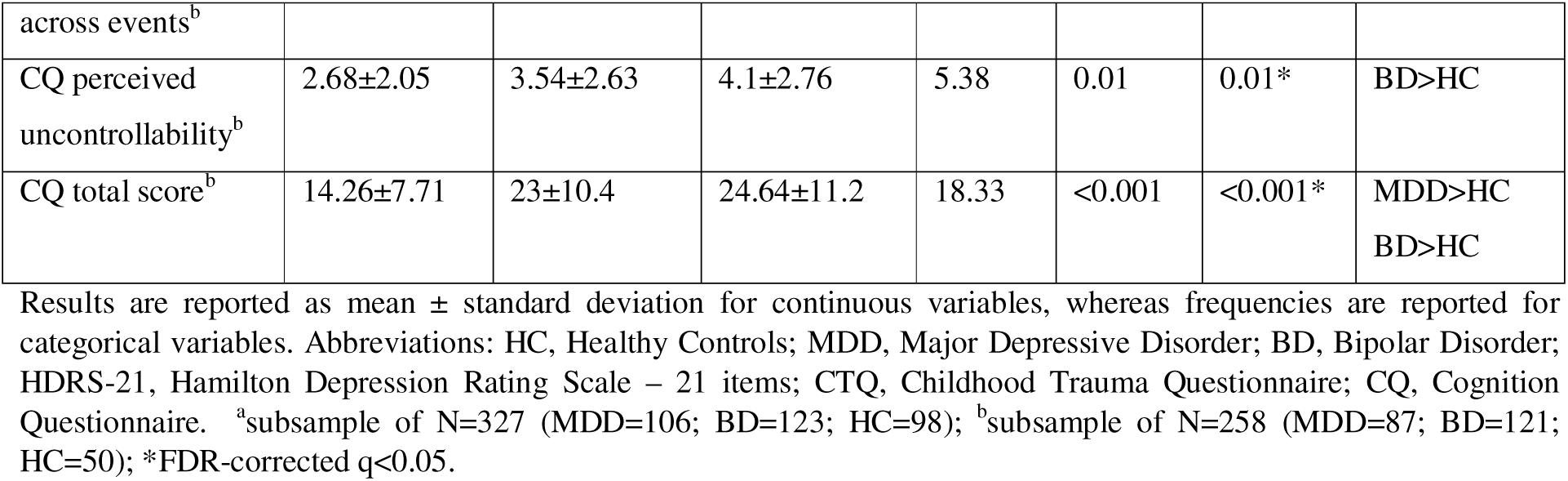
Descriptive statistics of the whole sample.

### Performances of stratification models

Models based on structural neuroimaging, FC or multimodal data showed good accuracies in clusters’ differentiation, identifying two clusters of patients as the optimal clustering solution (accuracy range: 73.4%-84.8%, q<0.05). Models trained exclusively on ReHo and fALFF modality did not achieve a predictive accuracy above chance level or significance (**Table 2** and **Figure S2**). The best clustering performance was observed for the FC-based model, showing an accuracy in discriminating clusters of 84.8%. Fleiss’ kappa statistic was k=0.07, suggesting a low agreement across stratification models in assigning subjects to the same cluster.

**Table 2.** Clustering models’ performance.

| Model | Algorithms | Clusters | Stability (error) | Acc | Sil | DB | q |
| --- | --- | --- | --- | --- | --- | --- | --- |
| FC | Kmeans + SVM | 2 | 0.15(0.02) | 84.8 | 0.29(2) | 1.24(2) | <0.001* |
| GM | Kmeans + SVM | 2 | 0.17(0.01) | 83.2 | 0.14(2) | 2.14(2) | <0.001* |
| CT | Kmeans + SVM | 2 | 0.18(0.02) | 81.9 | 0.27(2) | 1.51(2) | <0.001* |
| RD | Kmeans + SVM | 2 | 0.20(0.02) | 80.1 | 0.17(2) | 1.90(2) | 0.002* |
| FA | Kmeans + SVM | 2 | 0.21(0.01) | 79.2 | 0.20(2) | 1.73(2) | <0.001* |
| AD | Kmeans + SVM | 2 | 0.21(0.01) | 78.8 | 0.15(2) | 2.11(2) | <0.001* |
| MD | Kmeans + SVM | 2 | 0.24(0.02) | 76.5 | 0.20(2) | 1.84(2) | 0.04* |
| Multimodal | Multiview + SVM | 2 | 0.27(0.04) | 73.4 | 0.49(2) | 0.76(2) | 0.002* |
| ReHo | Kmeans + SVM | 2 | 0.59(0.04) | 40.9 | 0.07(2) | 3.21(3) | 0.044* |
| fALFF | Kmeans + SVM | 3 | 0.67(0.03) | 31.6 | 0.10(2) | 3.04(5) | 0.852 |

For each feature, number of clusters, normalized stability measures, and accuracies (in percentage) are reported. For comparison, the number of clusters identified based on internal measures (i.e., Silhouette and Davies-Bouldin scores) are reported in brackets. Models are sorted based on normalized stability measures, where lower values indicate a more stable clustering solution. FDR-corrected q-values obtained from 10,000 Monte-Carlo simulations are reported. Abbreviations: Acc, accuracy; Sil, Silhouette score; DB, Davies-Bouldin score; FC, functional connectivity; GM, grey matter; CT, cortical thickness; RD, radial diffusivity; FA, fractional anisotropy; AD, axial diffusivity; MD, mean diffusivity; ReHo, regional homogeneity; fALFF, fractional amplitude of low frequency fluctuations; SVM, support vector machine. \*FDR-corrected $q < 0.05$ .

### Neurobiological characterization of data-driven clusters

All the stratification models distinguished between a cluster of depressed patients with a more preserved brain structure or function and a second one characterized by a more compromised brain signature. Considering the FC-based clusters, Cluster 1 exhibited higher Spos compared to Cluster 2, with the largest effect sizes found in the cingulate gyrus, precuneus, and superior frontal gyrus (SFG). Conversely, Cluster 2 showed overall higher Sneg compared to Cluster 1, despite reduced effect sizes compared to Cluster 1 (**Figure 2**). Similarly, GM and CT-based models uncovered a cluster of depressed patients marked by widespread volumetric reductions and cortical thinning, with the main effects localized in temporo-parietal regions (**Figure S3**). All the DTI-based models uncovered one cluster differing from the second one in opposite direction, with the largest contributions in the corpus callosum and corona radiata (**Figure S4**). When combing all modalities in the multimodal stratification model, clusters’ differentiation was mainly guided by DTI measures, with one cluster exhibited higher RD, MD, AD, and lower FA values compared to the other (**Figure S5**).

To clarify the direction of neurobiological alterations in the best-performing stratification model (i.e., FC-based), we examined differences in global and regional Spos and Sneg measures between clusters and HC. At the global level, patients in Cluster 1 showed significantly higher Spos compared to both Cluster 2 and HC, while patients belonging to Cluster 2 exhibited elevated Sneg relative to Cluster 1 and HC (**Figure 3A-B**). Evidence of group differences in Spos or Sneg was also observed across individual brain regions, consistent with the patterns described at the global level (**Table S13**). These results generally indicate that Cluster 1 is distinguished by an increased functional integration, while an abnormal functional segregation characterize Cluster 2.

### Differential neurobiological-ACEs associations between clusters

A significant LV pair surviving FDR correction was obtained only for the FC-based model (explained covariance=40.0%, q=0.01) (**Table S11**). The largest BSR were observed for ROIs in the ventral and caudal part of the cingulate gyrus, superior and inferior parietal lobules, insula, somatosensory cortex, as well as the lateral amygdala, thalamus, and basal ganglia, and these effects were found for both Spos and Sneg (**Figure 4A-B**). In Cluster 1 all CTQ domains, except for sexual abuse, significantly contributed to the neurobiological-ACEs multivariate association. The direction of CTQ loadings was negative, meaning that higher exposure to childhood trauma is associated with decreased FC strength. In Cluster 2, negative significant associations emerged for emotional neglect, while positive for sexual abuse. (**Figure 4C**). PLS analysis including HC produced similar results, isolating a significant LV pair (explained covariance=43.0%, q=0.015) (**Table S12**). The contribution of CTQ domains to the neurobiological–ACEs association was highly similar between HC and Cluster 1, whereas only the domains of sexual and physical abuse were significant in Cluster 2. Thus, the identified LV likely reflects a normative neurobiological–ACEs relationship that is present in HC and preserved in Cluster 1, but absent in Cluster 2 (**Figure S6**).

When considering univariate analyses, patients belonging to the FC-based Cluster 2 showed higher scores in generalization across events and perceived uncontrollability, while a trend toward significance was found for sexual abuse. However, these results did not survive FDR correction (**Table S2**). Compared to HC, both clusters exhibited higher scores in attribution of causality, generalization across events, and CQ total score. However, only Cluster 2 showed higher scores in sexual abuse, CTQ total score, generalization across time, and perceived uncontrollability relative to HC (**Table S10**). At an uncorrected p-values, significant differences in diagnosis were found for clusters derived from AD, while a trend toward significance was detected in the FA-based clusters. No significant differences were detected for clusters derived from GM and CT (**Tables S3-S9**).

### Effect of FC strength on cognitive distortions in the FC-based clusters

The final step was to explore whether the LV FC scores differentially mediated the relationships between the LV CTQ scores and CQ domains in the FC-based clusters. Multivariate GLMs did not show significant effects for LV FC scores on CQ domains neither in Cluster 1 (Wilk’s λ=0.88, F(5,27)=0.73, p=0.612) nor in Cluster 2 (Wilk’s λ=0.87, F(5,58)=1.71, p=0.147). Considering univariate results, LV FC scores significantly and positively predicted generalization across time only in Cluster 2 (b=0.029, p=0.032; overall model fit: R2=0.07, RMSE=0.90, F(1,62)=4.85, p=0.031). Mediation analysis revealed a significant indirect effect of LV CTQ scores on generalization across time through LV FC scores (i.e., path a*b). The total and the direct effects of LV CTQ scores on generalization across time were not significant; however, the significant indirect effect support the mediation effect of LV FC scores. LV CTQ scores significantly increased LV FC scores (path a), whereas the effect of LV FC scores on generalization across time is marginally significant (path b). These results suggest that in Cluster 2 LV FC scores significantly mediate the relationship between LV CTQ scores and generalization across time, despite the total effect is not significant (**Figure 4D)**.

## Discussion

This study explored whether multimodal neuroimaging could support clinically meaningful data-driven stratifications of depressive mood disorders. Among all tested models, the highest accuracy was observed for the FC-based model (85%), uncovering two transdiagnostic biotypes: one characterized by widespread high positive FC strength (Cluster 1, N=50), while another one by dominant negative FC strength (Cluster 2, N=96). Importantly, differential neurobiological-ACEs associations in the FC-based clusters emerged. In Cluster 1, childhood trauma was negatively associated with FC in the cingulate gyrus, insula, somatosensory cortex, and subcortical regions, consistent with the ACEs-related pattern also observed in HC. In Cluster 2, emotional neglect was linked to reduced FC, while sexual abuse showed a positive association. ACEs-related FC mediated the relationship between ACEs and NCS of overgeneralization in this latter cluster, supporting biotype-specific neurobiological pathways linking early trauma and cognitive vulnerability.

The functional profiles of the discovered biotypes reflect widespread connectome alterations characterized by differential contribution of positive and negative FC strength. While positive correlations may reflect synchronized neural activity, the biological meaning of negative correlations is still debated (Goelman et al., 2014; Zhan et al., 2017). However, the strength of negative correlations among brain regions has been shown to be directly correlated with the length of their shortest path, thus reflecting less efficient information flow (Chen et al., 2011). The lack of balance between functional integration and segregation is well-documented in psychiatric disorders, possibly reflecting a shift toward a more random and less integrated network configuration (de Lange et al., 2019; Hansen et al., 2022; Repple et al., 2023; Xia et al., 2019). Our findings extend this scenario by demonstrating that this unbalance manifests as distinct biotypes within depression, differing from HC in global and regional FC.

A significant neurobiological-ACEs relationship was differentially present in the two FC biotypes. The regions most affected by ACEs (amygdala, cingulate, thalamus, basal ganglia, insula, sensorimotor areas), are crucial for emotional, cognitive and social functioning, and their functional organization is highly influenced by cortical maturational changes (Casey et al., 2019; Dong et al., 2024; Luo et al., 2024; Park et al., 2024). The deprivation of adequate emotional stimulation during childhood disrupts these processes by diminishing neurotrophin and neurotransmitters production which, ultimately, end with aberrant neural organization (De Bellis, 2005; van der Kolk, 2003). Among all the CTQ domains, sexual abuse emerged as a key discriminating factor between the two biotypes and HC, showing a significant and positive association with FC only in Cluster 2. Given the negative FC strength characterizing this biotype, exposure to childhood abuse would be expected to further reduce FC. However, this association may reflect a compensatory mechanism, consistent with the stress acceleration hypothesis, whereby early adversities accelerate the maturation of brain circuits involved in salience detection and executive functions to promote short-term adaptations (Callaghan and Tottenham, 2016; Holz et al., 2023). These findings may point to a more severely affected subgroup, where localized increases in functional coupling compensate for abuse-related widespread connectivity deficits.

The presence of distinct underlying mechanisms is further supported by the mediation analysis, where the ACE-related FC scores mediated the relationship between the ACEs and generalization across time only in the hyper-segregated biotype. Overgeneralization represents a key feature of depressive symptomatology (Gamble et al., 2019), and constitutes a core element of the depressive “cognitive triad” (Beck, 2008). While ACEs are established risk factors for NCS (Alaftar and Uzer, 2022; Mansueto et al., 2021; Poletti et al., 2014a), these results suggest that the tendency to overgeneralize might characterize a specific neurobiological subtype of depressed patients, which may benefit from cognitive or trauma-focused psychotherapies (Cohen and Mannarino, 2015; Wiles et al., 2013).

Unlike the FC-based biotypes, structural MRI models did not reveal significant associations with ACEs or NCS, though they reliably distinguished patients with widespread WM dysconnectivity, cortical thinning and volumetric reductions (accuracies > 70%). These alterations may reflect diagnosis-specific effects, consistent with evidence that GM and DTI metrics differentiate BD and MDD with accuracies up to 78% (Calesella et al., 2024; Colombo et al., 2022; Vai et al., 2020). Accordingly, clusters derived from FA and AD exhibited trends toward a significant difference for diagnosis, although not surviving FDR correction. Conversely, this association was not found for the FC-based clusters, suggesting that functional measures are more likely to reflect transdiagnostic signatures rather that clinical diagnoses (Goldstein-Piekarski et al., 2022; Tozzi et al., 2024; Williams, 2017). Alternatively, the limited overlap with FC clusters may reflect distinct pathological dimensions captured solely by structural modalities, such as immune-inflammatory mechanisms or cognitive deficits (Colombo et al., 2024; Liang et al., 2019). The low overlapping between structural and functional clustering solutions might also explain why the multimodal stratification model did not outperform the FC-based one, despite achieving a moderate classification performance (73.4%) and the best internal validity (Silhouette and Davies-Bouldin scores).

Some limitations should be underscored. All patients were inpatients admitted to the same hospital, limiting generalizability. Although pharmacological load was statistically controlled, residual long-lasting effects cannot be completely ruled out. Finally, the modest sample size and lack of additional biological measures (e.g., genetic or inflammatory markers) may have constrained detection of more nuanced associations.

In conclusion, this work shows that FC-based biotypes best captured distinct trauma-related neurobiological profiles in depression. One biotype exhibited hyperconnectivity and negative associations with ACEs; the other, characterized by network segregation, showed a more specific pattern involving sexual abuse, emotional neglect, and overgeneralization. These findings highlight the potential of FC to define transdiagnostic depression subtypes with differential vulnerability to early adversity, informing a neurobiologically grounded taxonomy of mood disorders.

## Data Availability

All data produced in the present study are available upon reasonable request to the authors

## Acknowledgments

None.

## Funding sources

This study was supported by the Italian Ministry of Health (grant numbers GR 2019-12370616).

---

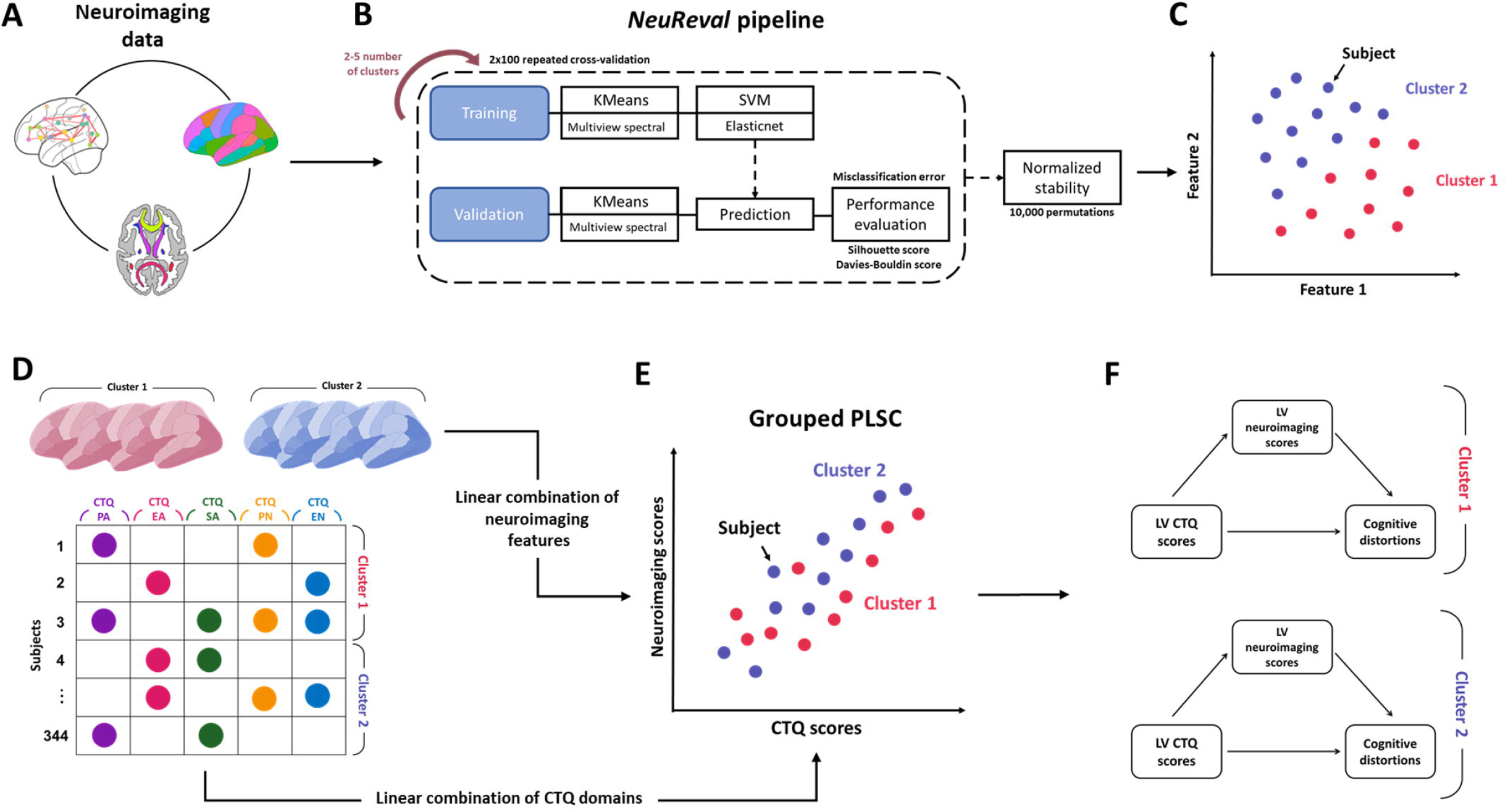

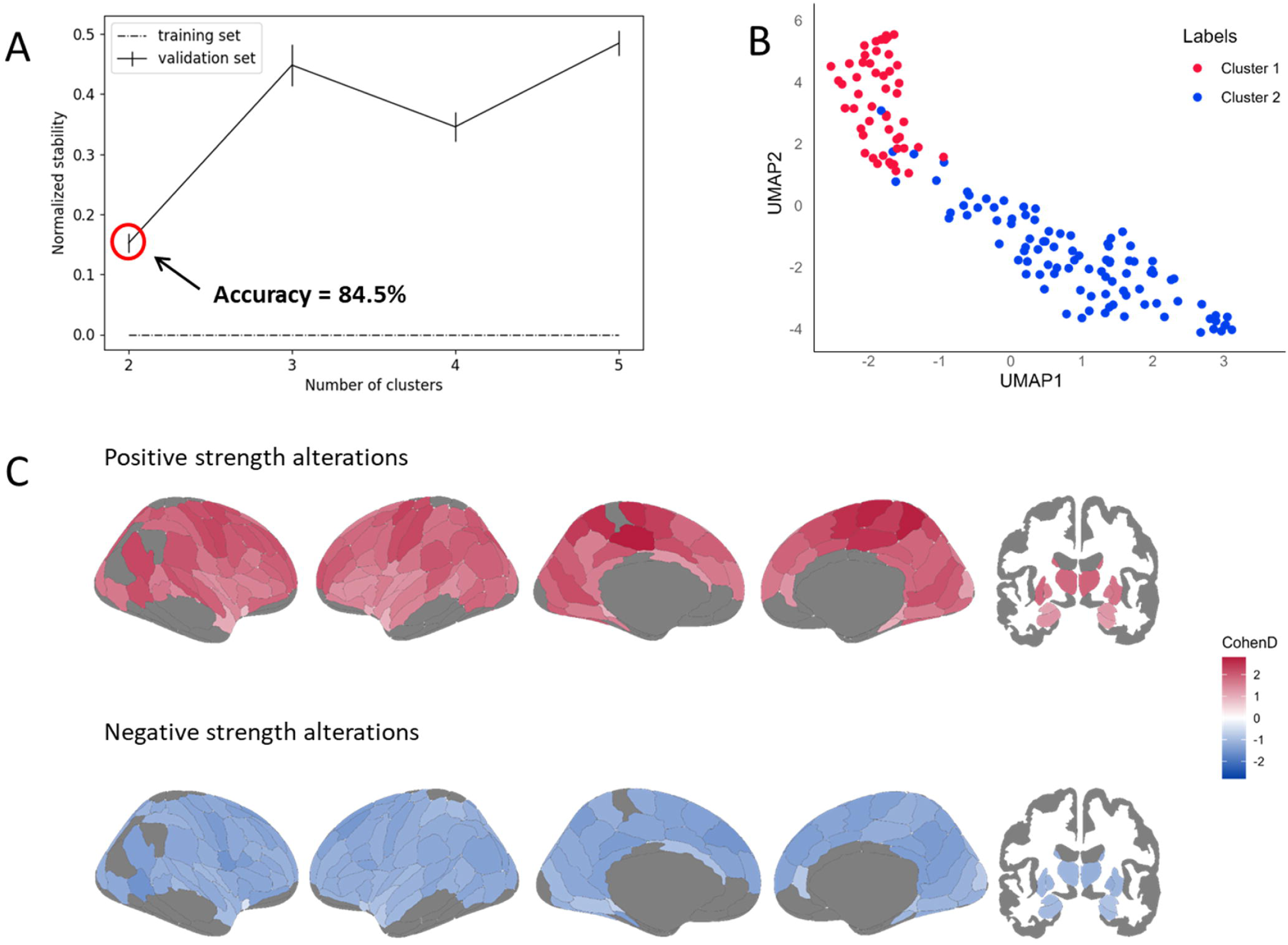

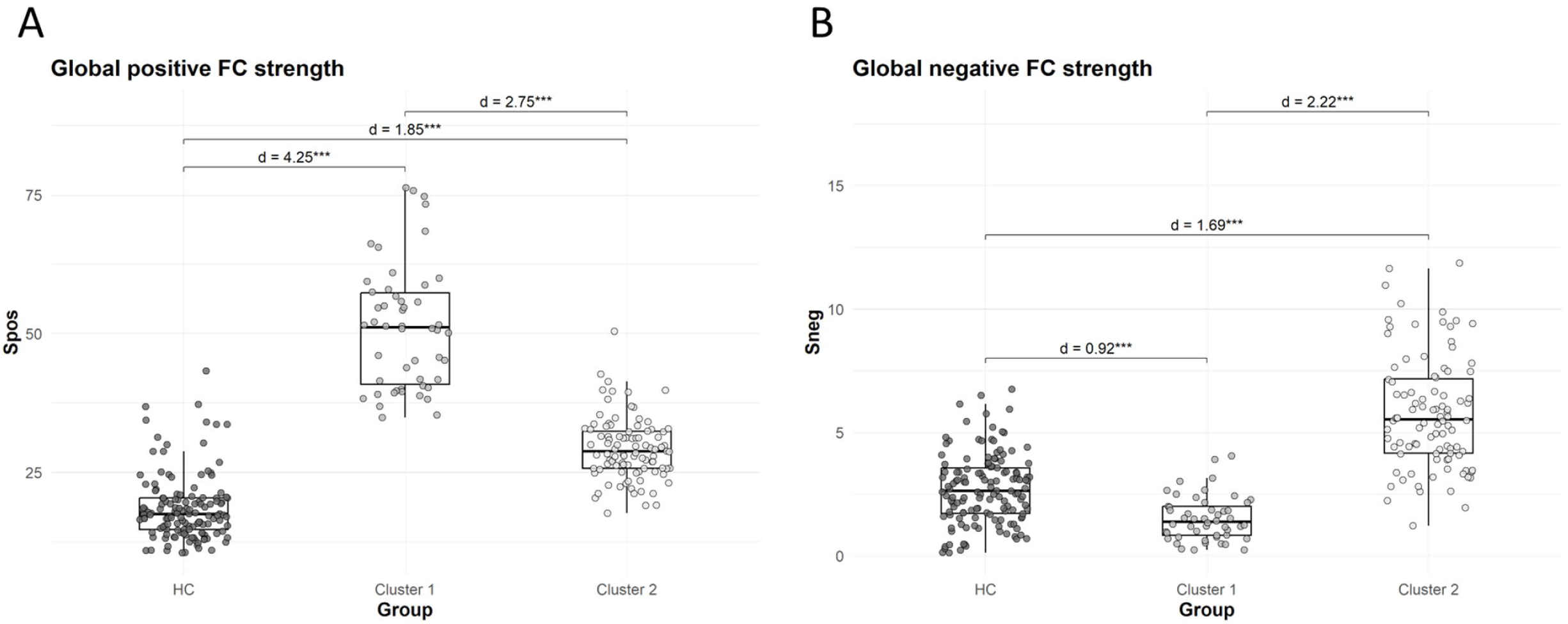

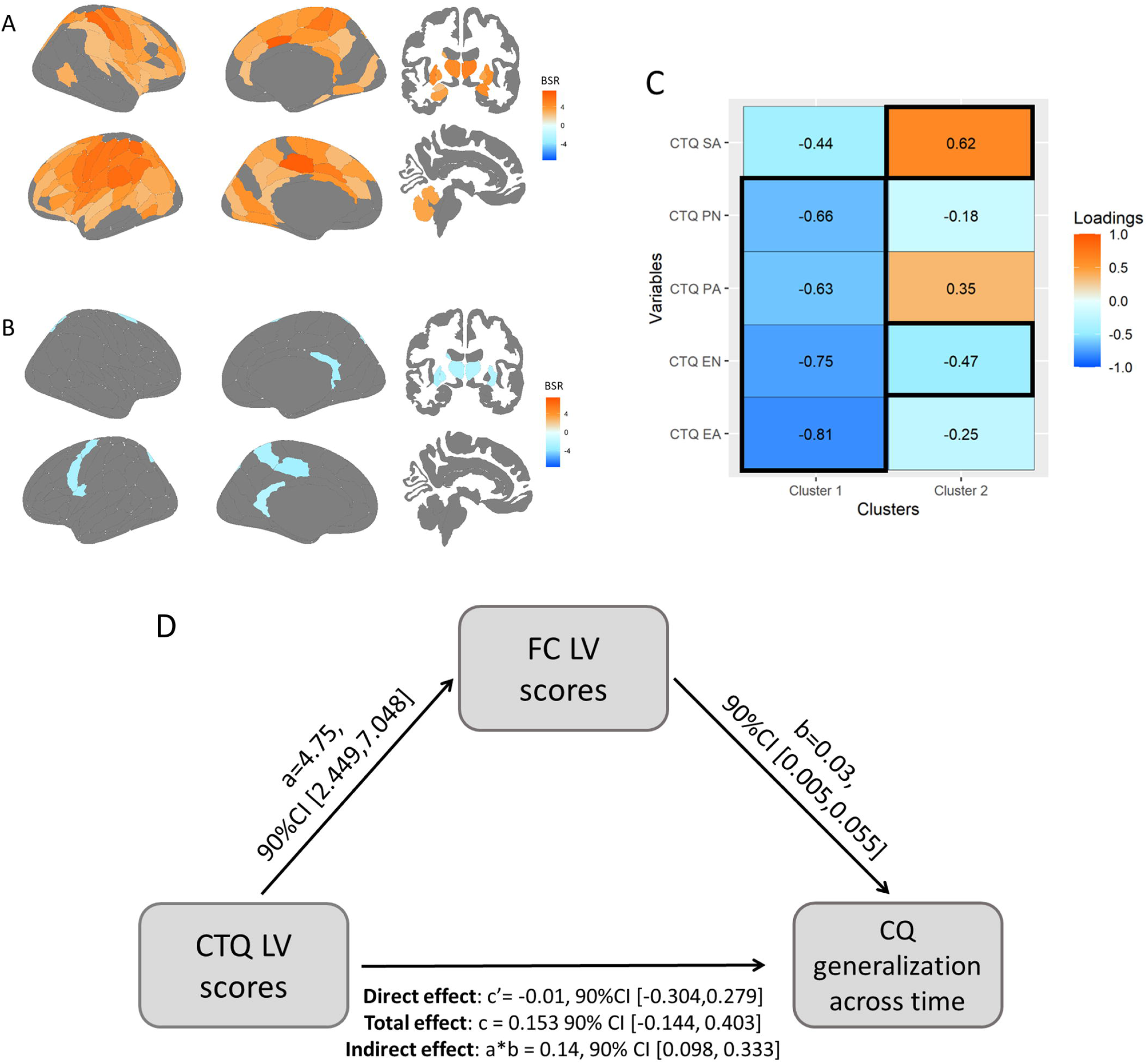

## Notes

### Competing Interest Statement

The authors have declared no competing interest.

### Author Declarations

Ethics Committee of IRCSS San Raffaele Scientific Institute (Milan, Italy) gave ethical approval for this work.

### Summary of Updates

Author affiliations updated; Figure 4 revised; discussion updated to clarifiy; declarations of interests updated.

