## Supplementary material for "Biotypes of deeply phenotyped depressed patients reflect signatures of adverse childhood experience and depressive cognitive biases": Methods S1

**Supplementary materials**

### *Methods S1. Rating criteria for medication load calculation*

To calculate a composite medication score for each patient, individual scores were assigned for each medication category, including: atypical antipsychotics, typical antipsychotics, benzodiazepines, mood stabilizers, serotonin-norepinephrine reuptake inhibitors (SNRI), selective serotonin reuptake inhibitors (SSRI), tricyclic antidepressant, anticonvulsants. These scores were then summed to obtain a patient-specific overall score. Mediation dosages were rated from low to high dosage according to specific criteria outlined by Sackeim (1). The following general guidelines were used for classification:

1. Any trial lasting less than 4 weeks, or monotherapies involving medications without established efficacy for unipolar depression, were assigned a score of “1”, regardless of dosage.
2. For alprazolam, specific anticonvulsants, and lithium, the maximum score was set at “2”.
3. Scores were reduced in case of lack of compliance and/or trial discontinuation.
4. In the case of combined therapies, each medication was rated independently. An exception was made for lithium: if taken for at least 2 weeks in combination with an antidepressant rated above “3”, the overall antidepressant score was increased by 1 point.

### *Methods S2. MRI acquisition parameters*

A sample of 192 patients (48 MDD, 144 BD) was acquired with Gyroscan Intera, Philips, Netherlands employing a 8 channels SENSE head coil (T1-weighted MPRAGE sequences: TR 25.00 ms, TE 4.6 ms, field of view FOV=230 mm,91 matrix=256×256, in-plane resolution 0.9×0.9 mm, yielding 220 transversal slices with a thickness of 0.8 mm). SE EPI sequences (TR/TE=9000/58 ms, FoV (mm) 232(ap), 126 (fh), 240.00 (rl); acquisition matrix = 112×85; voxel acquisition 2.14×2.73×2.3; 55 contiguous, with in-plane voxel size 1.88×1.88 mm; SENSE acceleration factor=2; 1 b0 and 35 non-collinear directions of the diffusion gradients; b value=900 s/mm2) were used. Finally, for 50 patients (22 MDD, 28 BD), a resting-state fMRI acquisition was also performed, with the instruction to keep their eyes closed. Gradient-echo EPI sequences were acquired. The scanning sessions included 100 sequential T2*-weighted volumes (interleaved ascending transverse slices covering the whole brain), acquired using an EPI pulse sequence (TR = 2000 ms; TE = 30 ms; flip angle = 77°; field of view = 220 mm; number of slices = 32; slice thickness = 4 mm; matrix size = 64 x 63 reconstructed up to 64 x 64 pixels). Two dummy scans before fMRI acquisition allowed for obtaining longitudinal magnetization equilibrium. Total time acquisition was 3 min and 28 s.

A sample of 152 patients (87 MDD, 65 BD) and 138 HC was instead acquired with an Ingenia CX, Philips, The Netherlands using a 32-channel sensitivity encoding SENSE head coil (T1-weighted MPRAGE sequence: TR 8.00 ms, TE 3.7 ms, field of view FOV = 256 mm, matrix = 256 x 256, in-plane resolution 1 x 1 mm, yielding 182 transversal slices with a thickness of 1 mm). SE EPI sequences (EPI factor= 43; TR/TE=5900/78 ms, FoV (mm) 232 (ap), 129 (fh), 240.00 (rl); acquisition matrix 112×85; 56 contiguous, 2.3-mm thick axial slices reconstructed with in-plane pixel size 1.88×1.88 mm; SENSE acceleration factor= 2; Multiband acceleration factor= 2; ten b0 and 96 non-collinear directions of the diffusion gradients: 60 b values=2855 s/mm2, 6 b values=700 s/mm2, 30 b values=1000 s/mm2) were acquired. Fat saturation was performed to avoid chemical shift artefacts. Lastly, for 96 patients (57 MDD, 39 BD) and 138 HC, a resting-state fMRI acquisition was performed, with the instruction to keep their eyes closed. Gradient-echo EPI sequences were acquired. The scanning sessions comprised 200 sequential T2*-weighted volumes (interleaved ascending transverse slices covering the whole brain, tilted 30° downward concerning bicommissural line to reduce susceptibility artefacts in orbitofrontal region), acquired using an EPI pulse sequence (TR = 2000 ms; TE = 30 ms; flip angle = 85°; field of view = 192 mm; number of slices = 38; slice thickness = 3.7 mm; matrix size = 64 x 62 reconstructed up to 96 x 96 pixels). Six dummy scans before fMRI acquisition allowed for obtaining longitudinal magnetization equilibrium. Total time acquisition was 6 min and 56 s.

### *Figure S1. Correlations between CTQ and CQ domains in the whole sample*

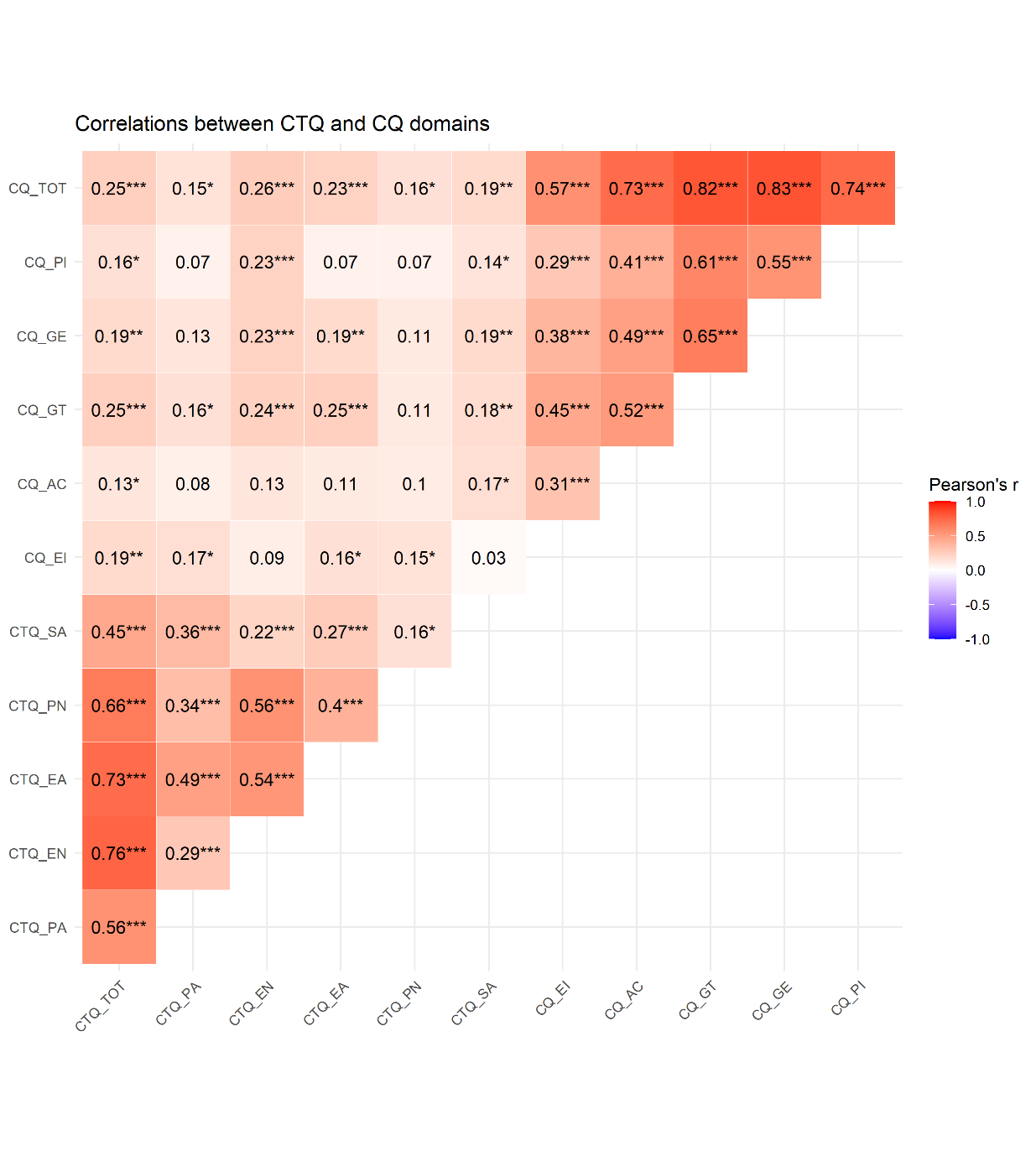

Asterisks indicate statistically significant correlations that survive p<0.05. Abbreviations: CTQ, Childhood Trauma Questionnaire; CQ, Cognition Questionnaire; SA, sexual abuse; PN, physical neglect; PA, physical abuse; EN, emotional neglect; EA, emotional abuse; EI, emotional impact; AC, attribution of causality; GT, generalization across time; GE, generalization across events; PI, perceived uncontrollability.

.

### *Figure S2. Normalized stability plots for each clustering model*

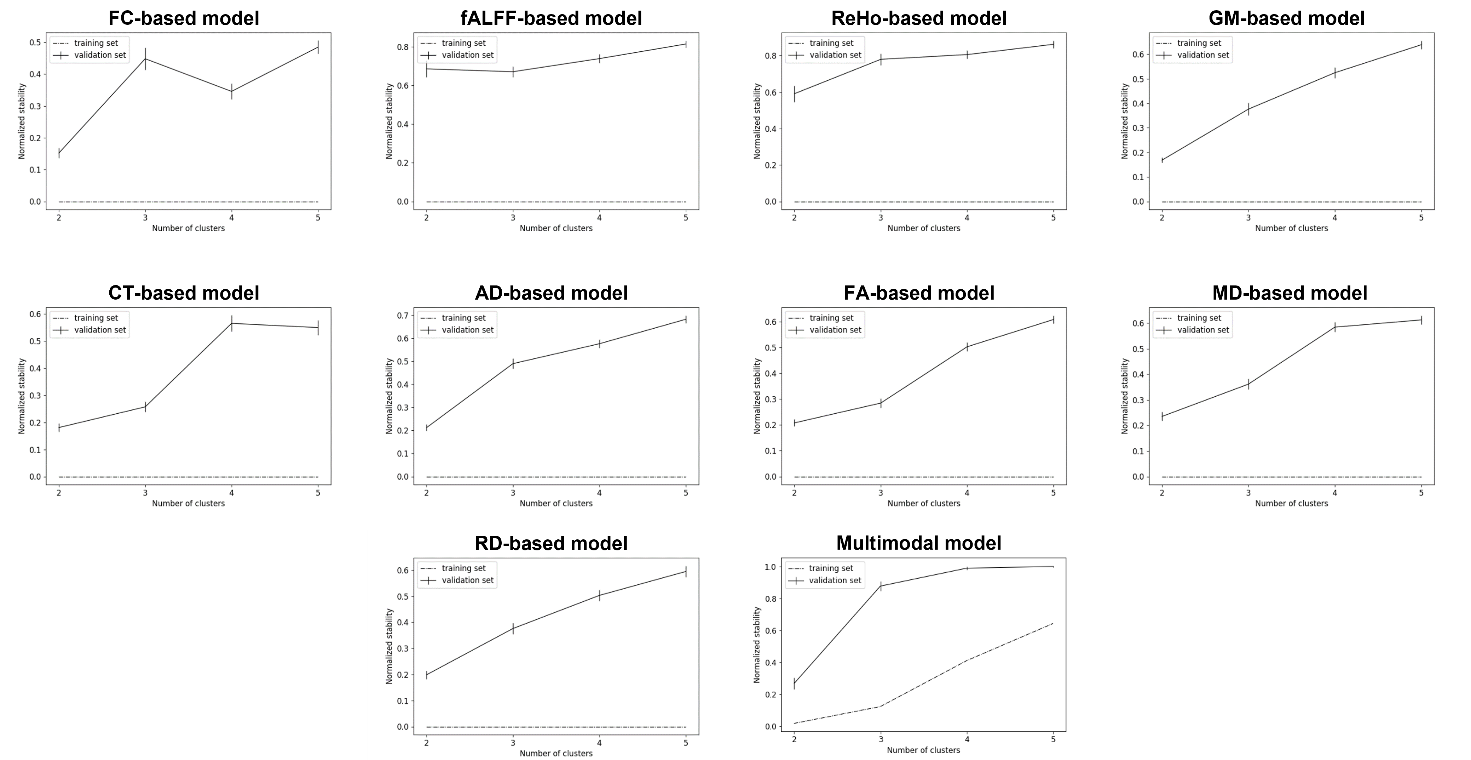

Solid lines represent the validation normalized stability with 95% confidence intervals. Dashed lines indicate the stability during training. Abbreviations: FC, functional connectivity; fALFF, fractional amplitude of low frequency fluctuations; ReHo, regional homogeneity; GM, grey matter; CT, cortical thickness; AD, axial diffusivity; FA, fractional anisotropy; MD, mean diffusivity; RD, radial diffusivity.

### *Figure S3. Results of stability-based relative clustering validation on grey matter volumes and cortical thickness.*

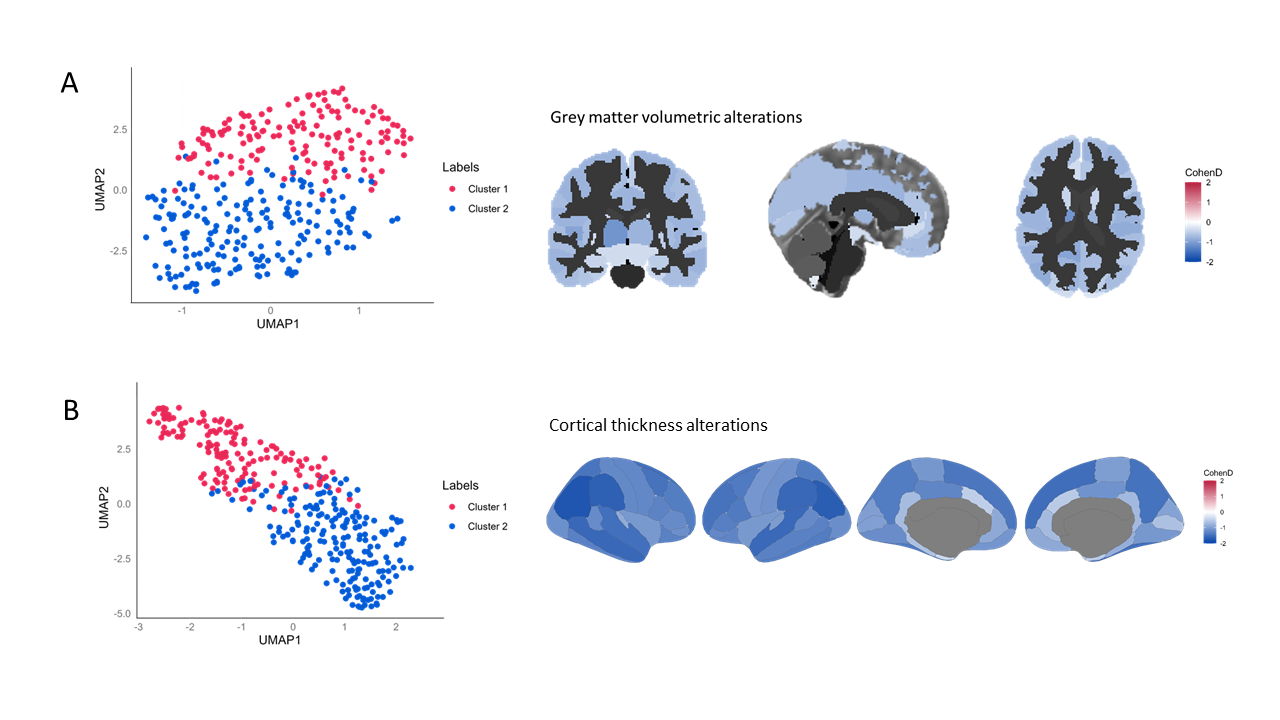

A. UMAP plot (left) and Cohen's d values describing the magnitude of differences in regional grey matter volumes between clusters (right). B. UMAP plot (left) and Cohen's d values describing the magnitude of differences in cortical thickness between clusters (right). Positive values (red) indicate higher values for Cluster 1 compared to Cluster 2, whereas negative values (blue) indicate higher values for Cluster 2 compared to Cluster 1. Abbreviations: UMAP, Uniform Manifold Approximation and Projection for Dimension Reduction.

### *Figure S4. Results of stability-based relative clustering validation on DTI measures.*

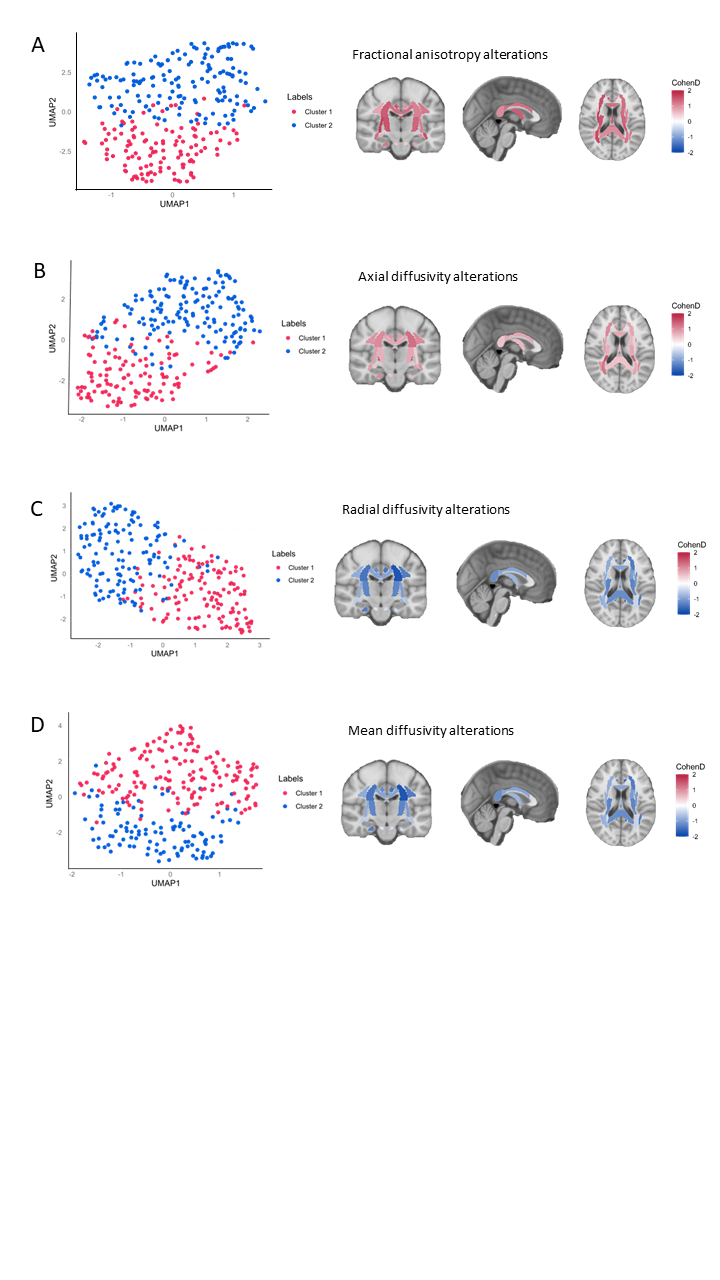

For each DTI model, UMAP plots (left) and Cohen's d values (right) describing the magnitude of differences in fractional anisotropy (A), axial (B), radial (C), and mean diffusivity (D) between clusters are reported. Positive values (red) indicate higher values for Cluster 1 compared to Cluster 2, whereas negative values (blue) indicate higher values for Cluster 2 compared to Cluster 1. Abbreviations: UMAP, Uniform Manifold Approximation and Projection for Dimension Reduction.

### *Figure S5. Results of the multimodal stability-based relative clustering validation model.*

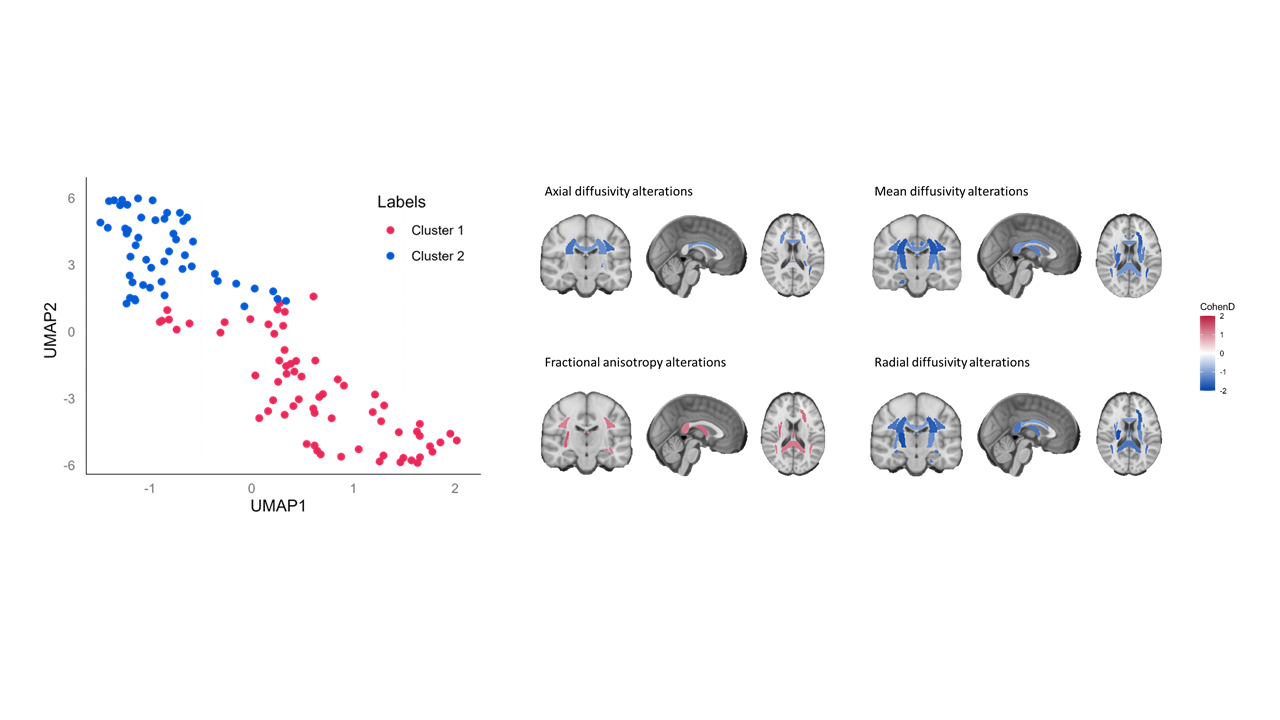

UMAP plots (left) and Cohen's d values (right) describing the magnitude of differences in DTI measures between clusters. Positive values (red) indicate higher values for Cluster 1 compared to Cluster 2, whereas negative values (blue) indicate higher values for Cluster 2 compared to Cluster 1. Abbreviations: UMAP, Uniform Manifold Approximation and Projection for Dimension Reduction.

### *Figure S6. Differences in multivariate brain-ACEs relationships between the FC-based clusters and healthy controls.*

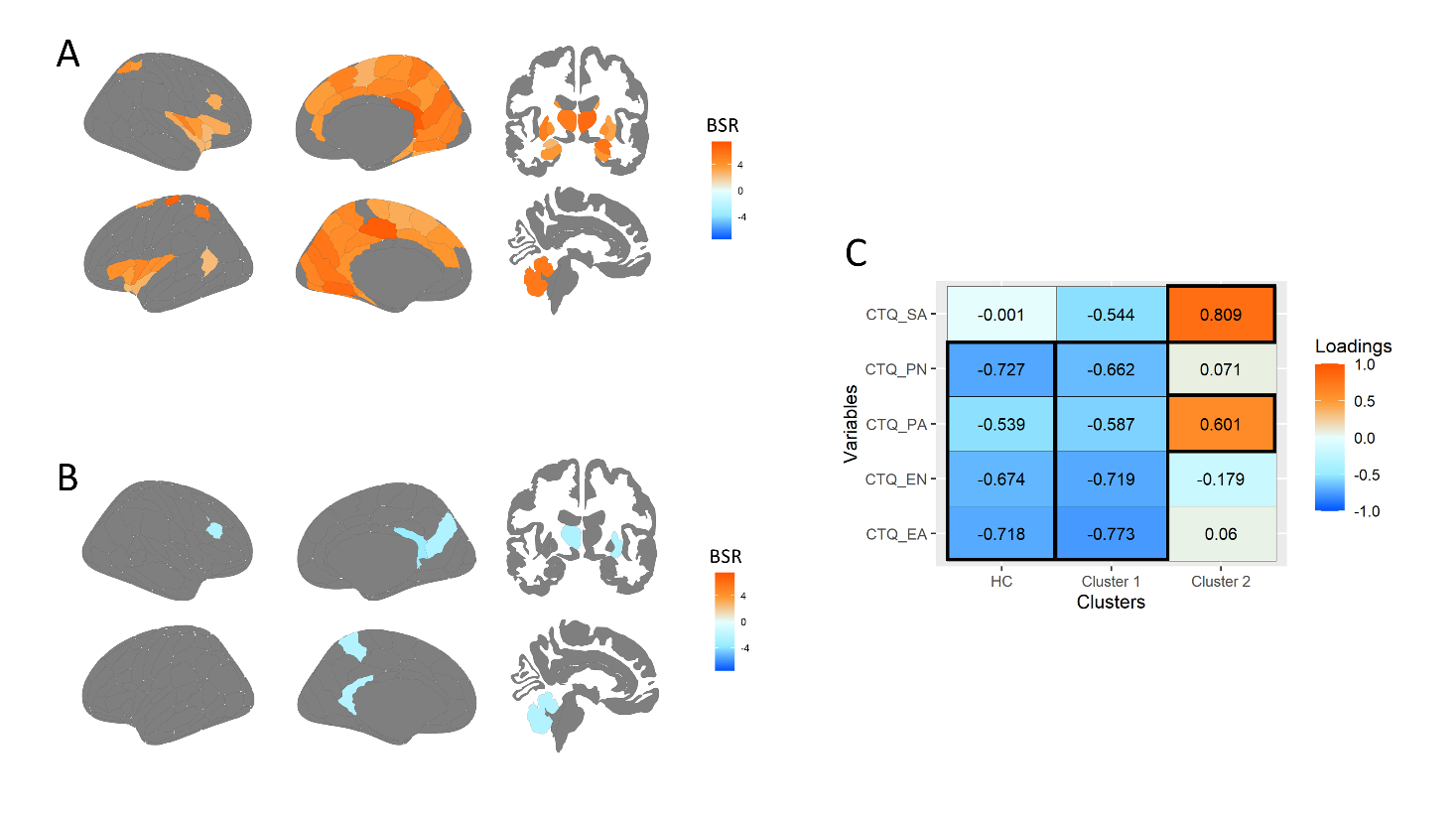

A-B. Significant brain bootstrap ratios (BSRs) for positive (A) and negative (B) FC strength in each region. Brain regions with orange BSRs have correlations that have the same directionality shown in the heatmap in (C). Brain regions with blue BSRs have correlations in the opposite direction shown in the heatmap in (C). C. PLS correlations for each CTQ domain (row) and each cluster (columns). Loadings with black outline are those with 95% confidence intervals that do not include zero. Abbreviations: BSR, brain bootstrap ratio; CTQ, Childhood Trauma Questionnaire; SA, sexual abuse; PN, physical neglect; PA, physical abuse; EN, emotional neglect; EA, emotional abuse.

### *Table S1. List of the selected ROIs for fMRI features*

| **Brainnettome label** | **ROI label** |
| --- | --- |
| A8m L | SFG medial frontal area 8 L |
| A8m R | SFG medial frontal area 8 R |
| A8dl L | SFG dorsolateral frontal area 8 L |
| A8dl R | SFG dorsolateral frontal area 8 R |
| A9l L | SFG lateral area 9 L |
| A9l R | SFG lateral area 9 R |
| A6dl L | SFG dorsolateral area 6 L |
| A6dl R | SFG dorsolateral area 6 R |
| A6m L | SFG medial area 6 L |
| A6m R | SFG medial area 6 R |
| A9m L | SFG medial area 9 L |
| A9m R | SFG medial area 9 R |
| A10m L | SFG medial area 10 L |
| A10m R | SFG medial area 10 R |
| A9/46d L | MFG dorsal area 9/46 L |
| A9/46d R | MFG dorsal area 9/46 R |
| IFJ L | MFG inferior frontal junction L |
| IFJ R | MFG inferior frontal junction R |
| A46 L | MFG middle frontal area 46 L |
| A46 R | MFG middle frontal area 46 L |
| A9/46v L | MFG ventral area 9/46 L |
| A9/46v R | MFG ventral area 9/46 R |
| A8vl L | MFG ventrolateral area 8 L |
| A8vl R | MFG ventrolateral area 8 R |
| A6vl L | MFG ventrolateral area 6 L |
| A6vl R | MFG ventrolateral area 6 R |
| A44d L | IFG dorsolateral area 44 L |
| A44d R | IFG dorsolateral area 44 L |
| IFS L | IFG inferior frontal sulcus L |
| IFS R | IFG inferior frontal sulcus R |
| A45c L | IFG caudal area 45 L |
| A45c R | IFG caudal area 45 R |
| A45r L | IFG rostral area 45 L |
| A45r R | IFG rostral area 45 R |
| A44op L | IFG opercular area 44 L |
| A44op R | IFG opercular area 44 R |
| A44v L | IFG ventral area 44 L |
| A44v R | IFG ventral area 44 L |
| A12/47l L | OrG lateral area 12/47 L |
| A12/47l R | OrG lateral area 12/47 R |
| A4hf L | PreCG area 4 head and face L |
| A4hf R | PreCG area 4 head and face R |
| A6cdl L | PreCG caudal dorsolat area 6 L |
| A6cdl R | PreCG caudal dorsolat area 6 R |
| A4ul L | PreCG area 4 upper limb 4 L |
| A4ul R | PreCG area 4 upper limb 4 R |
| A4tl L | PreCG area 4 tongue and larynx L |
| A4tl R | PreCG area 4 tongue and larynx R |
| A6cvl L | PreCG caudal ventrolateral area 6 L |
| A6cvl R | PreCG caudal ventrolateral area 6 R |
| A1/2/3ll R | ParaCL area 1/2/3 lower limb R |
| A4ll L | ParaCL area 4 lower limb L |
| A4ll R | ParaCL area 4 lower limb R |
| A41/42 L | STG area 41/42 L |
| A41/42 R | STG area 41/42 R |
| TE1.0/TE1.2 L | STG TE 1.0/1.2 L |
| TE1.0/TE1.2 R | STG TE 1.0/1.2 R |
| A22c L | STG caudal area 22 L |
| A22c R | STG caudal area 22 R |
| A38l L | STG lateral area 38 L |
| A38l R | STG lateral area 38 R |
| A22r L | STG rostral area 22 L |
| A22r R | STG rostral area 22 R |
| A37dl L | MTG dorsolateral area 37 L |
| A37dl R | MTG dorsolateral area 37 R |
| A37vl L | ITG ventrolateral area 37 L |
| A37mv L | FusGy medioventral area 37 L |
| A37mv R | FusGy medioventral area 37 R |
| TL L | ParHipGy lateral post PH area L |
| TL R | ParHipGy lateral post PH area R |
| TH L | ParHipGy Tharea medialPHG L |
| TH R | ParHipGy Tharea medialPHG R |
| rpSTS L | PostSupTempSulc rostropost sup temp sulc L |
| rpSTS R | PostSupTempSulc rostropost sup temp sulc R |
| cpSTS L | PostSupTempSulc caudopost sup temp sulc L |
| cpSTS R | PostSupTempSulc caudopost sup temp sulc R |
| A7r L | SPL rostral area 7 L |
| A7r R | SPL rostral area 7 R |
| A7c L | SPL caudal area 7 L |
| A7c R | SPL caudal area 7 R |
| A5l L | SPL lateral area 5 L |
| A5l R | SPL lateral area 5 R |
| A7ip L | SPL intrapariet area 7 L |
| A7ip R | SPL intrapariet area 7 R |
| A39c L | IPL caudal area 39 L |
| A39rd L | IPL rostrodorsal area 39 L |
| A40rd L | IPL rostrodorsal area 40 L |
| A40rd R | IPL rostrodorsal area 40 R |
| A40c L | IPL caudal area 40 L |
| A39rv L | IPL rostroventral area 39 L |
| A39rv R | IPL rostroventral area 39 R |
| A40rv L | IPL rostroventral area 40 L |
| A40rv R | IPL rostroventral area 40 R |
| A7m L | PreCun medial area 7 L |
| A7m R | PreCun medial area 7 R |
| A5m L | PreCun medial area 5 L |
| A5m R | PreCun medial area 5 R |
| dmPOS L | PreCun dorsomed parietocc sulcus L |
| dmPOS R | PreCun dorsomed parietocc sulcus R |
| A31 L | PreCun area 31 L |
| A31 R | PreCun area 31 R |
| A1/2/3ulhf L | PostCGy area 1/2/3 head and face L |
| A1/2/3ulhf R | PostCGy area 1/2/3 head and face R |
| A1/2/3tonIa L | PostCGy area 1/2/3 tongue and larynx L |
| A1/2/3tonIa R | PostCGy area 1/2/3 tongue and larynx R |
| A2 L | PostCGy lateral area 2 L |
| A2 R | PostCGy lateral area 2 R |
| G L | InsL hypergranular insular L |
| G R | InsL hypergranular insular R |
| vIa L | InsL ventral agranular insular L |
| vIa R | InsL ventral agranular insular R |
| dIa L | InsL dorsolat agranular insular L |
| dIa R | InsL dorsolat agranular insular R |
| vId/vIg L | InsL ventral granular insular L |
| vId/vIg R | InsL ventral granular insular R |
| dIg L | InsL dorsolat granular insular L |
| dIg R | InsL dorsolat granular insular R |
| dId L | InsL dorsal dysgranular insular L |
| dId R | InsL dorsal dysgranular insular R |
| A23d L | CingGy dorsal area 23 L |
| A23d R | CingGy dorsal area 23 R |
| A24rv L | CingGy rostroventral area 24 L |
| A32p L | CingGy pregenual area 32 L |
| A32p R | CingGy pregenual area 32 R |
| A23v L | CingGy ventral area 23 L |
| A23v R | CingGy ventral area 23 R |
| A24cd L | CingGy caudodorsal area 24 L |
| A24cd R | CingGy caudodorsal area 24 R |
| A23c L | CingGy caudal area 23 L |
| A23c R | CingGy caudal area 23 R |
| A32sg R | CingGy subgenual area 32 R |
| cLinG L | MedVentOC caudal lingual gyrus L |
| cLinG R | MedVentOC caudal lingual gyrus R |
| rCunG L | MedVentOC rostral cuneus gyrus L |
| rCunG R | MedVentOC rostral cuneus gyrus R |
| cCunG R | MedVentOC caudal cuneus gyrus R |
| rLinG L | MedVentOC rostral lingual gyrus L |
| rLinG R | MedVentOC rostral lingual gyrus R |
| vmPOS L | MedVentOC ventromed parietocc sulcus L |
| vmPOS R | MedVentOC ventromed parietocc sulcus R |
| mOccG L | LatOC middle occip gyrus L |
| mOccG R | LatOC middle occip gyrus R |
| V5/MT+ L | LatOC V5/MT area L |
| V5/MT+ R | LatOC V5/MT area R |
| msOccG L | LatOC med sup occ gyrus L |
| msOccG R | LatOC med sup occ gyrus R |
| lsOccG L | LatOC lat sup occ gyrus L |
| lsOccG R | LatOC lat sup occ gyrus R |
| lAmyg L | Amy lateral amygdala L |
| lAmyg R | Amy lateral amygdala R |
| rHipp L | Hipp rostral hipp L |
| rHipp R | Hipp rostral hipp R |
| cHipp L | Hipp caudal hipp L |
| cHipp R | Hipp caudal hipp R |
| vCa L | BasGang ventral caudate L |
| vCa R | BasGang ventral caudate R |
| GP L | BasGang globus pallidus L |
| GP R | BasGang globus pallidus R |
| NAC L | BasGang nucleus accumbens L |
| vmPu L | BasGang ventromedial putamen L |
| vmPu R | BasGang ventromedial putamen R |
| dCa L | BasGang dorsal caudate L |
| dCa R | BasGang dorsal caudate R |
| dlPu L | BasGang dorsolat putamen L |
| dlPu R | BasGang dorsolat putamen R |
| mPFtha L | Thal medial prefront thalamus L |
| mPFtha R | Thal medial prefront thalamus R |
| mPMtha L | Thal premotor thalamus L |
| mPMtha R | Thal premotor thalamus R |
| Stha L | Thal sensory thalamus L |
| Stha R | Thal sensory thalamus R |
| rTtha L | Thal rostral temp thalamus L |
| rTtha R | Thal rostral temp thalamus R |
| PPtha L | Thal posterior parietal thalamus L |
| PPtha R | Thal posterior parietal thalamus R |
| Otha L | Thal occipital thalamus L |
| Otha R | Thal occipital thalamus R |
| cTtha L | Thal caudal temporal thalamus L |
| cTtha R | Thal caudal temporal thalamus R |
| lPFtha L | Thal lateral prefrontal thalamus L |
| lPFtha R | Thal lateral prefrontal thalamus R |
| Buckner2011 17Networks 1 | Buckner cerebellar networks 1 |
| Buckner2011 17Networks 2 | Buckner cerebellar networks 2 |
| Buckner2011 17Networks 4 | Buckner cerebellar networks 4 |
| Buckner2011 17Networks 5 | Buckner cerebellar networks 5 |
| Buckner2011 17Networks 9 | Buckner cerebellar networks 9 |

For each Brainnetome label the corresponding region of interest (ROI) is reported Abbreviations: SFG, superior frontal gyrus; MFG, medial frontal gyrus; IFG; inferior frontal gyrus; OrG, orbital gyrus; PreCG, precentral gyrus; ParaCL, paracentral lobule; STG, superior temporal gyrus; MTG, medial temporal gyrus; ITG, inferior temporal gyrus; FusGy, fusiform gyrus; ParHipGy, hippocampal part of the cingulate gyrus; PostSupTempSulc, posterior and superior temporal sulcus; SPL, superior parietal lobule; IPL, inferior parietal lobule; PreCun, precuneus; PostCGy, posterior cingulate gyrus; Ins, insula; CingCy, cingulate gyrus; MedVenOC, medial ventral occipital cortex; LatOC, lateral occipital cortex; Amy, amygdala; Hipp, hippocampus; BasGang, basal ganglia; Thal, thalamus; L, left; R, right.

### *Table S2. Clinical and demographic comparisons for the FC-based clusters*

| **Variable** | **Cluster 1 (N=50)** | **Cluster 2 (N=96)** | **t / U / χ2** | **p** | **q** |
| --- | --- | --- | --- | --- | --- |
| Age | 47.67 ± 11.57 | 47.67 ± 11.57 | 2329 | 0.77 | 0.91 |
| Sex | 35 F; 15 M | 64 F; 32 M | 0.05 | 0.82 | 0.91 |
| Diagnosis | 27 MDD; 23 BD | 52 MDD; 44 BD | 0 | 1.00 | 1.00 |
| Scanner | 16 Gyroscan; 34 Igenia | 34 Gyroscan; 16 Igenia | 0.05 | 0.82 | 0.91 |
| No. of episodes | 7.98 ± 10.21 | 9.47 ± 11.49 | 2087 | 0.19 | 0.52 |
| Age at onset | 30.36 ± 10.72 | 29.2 ± 10.43 | 2541.5 | 0.56 | 0.91 |
| Duration of illness (years) | 17.14 ± 10.71 | 18.47 ± 11.09 | 2235.5 | 0.50 | 0.91 |
| Years of education | 12.98 ± 3.236 | 12.74 ± 3.69 | 2511 | 0.63 | 0.91 |
| Pharmacological load | 4.54 ± 2.05 | 4.89 ± 2.26 | 2287.5 | 0.64 | 0.91 |
| HDRS-21 total score | 21.96 ± 5.98 | 21.49 ± 6.168 | 2602 | 0.40 | 0.88 |
| CTQ physical abuse^a^ | 5.82 ± 1.99 | 6.23 ± 2.68 | 1436 | 0.30 | 0.73 |
| CTQ emotional abuse^a^ | 8.33± 4.25 | 9.05 ± 3.93 | 1335 | 0.14 | 0.51 |
| CTQ sexual abuse^a^ | 5.44 ± 1.96 | 6.22 ± 3.15 | 1350.5 | 0.05 | 0.33 |
| CTQ physical neglect^a^ | 7.18 ± 2.92 | 7.31 ± 3.29 | 1604 | 0.98 | 1.00 |
| CTQ emotional neglect^a^ | 12.15 ± 4.53 | 12.68 ± 5.19 | 1524 | 0.68 | 0.91 |
| CTQ total score^a^ | 41.31 ± 13.25 | 43.17 ± 15.54 | 1495.5 | 0.57 | 0.91 |
| CQ emotional impact^b^ | 4.51 ± 2.71 | 4.28 ± 2.78 | 1264 | 0.70 | 0.91 |
| CQ attribution of causality^b^ | 5.06 ± 2.22 | 5.59 ± 2.23 | 1008 | 0.17 | 0.52 |
| CQ generalization across time^b^ | 3.89 ± 2.89 | 4.73 ± 2.86 | 968 | 0.10 | 0.44 |
| CQ generalization across events^b^ | 5.37 ± 2.92 | 6.59 ± 2.76 | -2.06 | **0.04** | 0.33 |
| CQ perceived uncontrollability^b^ | 2.97 ± 2.70 | 4.232 ± 2.691 | 867 | **0.02** | 0.33 |
| CQ total score^b^ | 21.69 ± 10.70 | 25.42 ± 10.20 | 934.5 | **0.06** | 0.33 |

Results are reported as mean ± standard deviation for continuous variables, whereas frequencies are reported for categorical variables. Significant differences are reported in bold. Abbreviations: MDD, Major Depressive Disorder; BD, Bipolar Disorder; HDRS-21, Hamilton Depression Rating Scale – 21 items; CTQ, Childhood Trauma Questionnaire; CQ, Cognition Questionnaire. ^a^subsample of N=121 (39 Cluster 1, 82 Cluster 2); ^b^subsample of N=104 (35 Cluster 1, 69 Cluster 2). *FDR-corrected q<0.05.

### *Table S3. Clinical and demographic comparisons for clusters derived from grey matter volumes.*

| **Variable** | **Cluster 1 (N=150)** | **Cluster 2 (N=194)** | **t / U / χ2** | **p** | **q** |
| --- | --- | --- | --- | --- | --- |
| Age | 48.29 ± 10.01 | 48.08 ± 10.68 | 14476 | 0.94 | 0.98 |
| Sex | 93 F; 57 M | 135 F; 59 M | 1.85 | 0.17 | 0.98 |
| Diagnosis | 65 MDD; 85 BD | 70 MDD; 124 BD | 1.57 | 0.21 | 0.77 |
| Scanner | 74 Gyroscan; 76 Igenia | 118 Gyroscan; 76 Igenia | 4.08 | **0.04** | 0.77 |
| No. of episodes | 9.073 ± 10.81 | 10.22 ± 11.61 | 13820.5 | 0.42 | 0.77 |
| Age at onset | 30.86 ± 10.48 | 30.31 ± 10.91 | 15085.5 | 0.56 | 0.98 |
| Duration of illness (years) | 17.43 ± 10.2 | 17.77 ± 11.26 | 14531.5 | 0.98 | 0.98 |
| Years of education | 12.63 ± 3.998 | 11.99 ± 3.73 | 15741 | 0.15 | 0.98 |
| Pharmacological load | 4.67 ± 2.34 | 4.54 ± 2.12 | 15214.5 | 0.46 | 0.77 |
| HDRS-21 total score | 22.35 ± 6.11 | 22.53 ± 5.52 | 14411.5 | 0.88 | 0.98 |
| CTQ physical abuse^a^ | 6.07 ± 1.87 | 6.37 ± 4.25 | 6828.5 | 0.50 | 0.77 |
| CTQ emotional abuse^a^ | 8.91 ± 4.15 | 8.17 ± 3.61 | 7162 | 0.20 | 0.77 |
| CTQ sexual abuse^a^ | 5.81 ± 2.26 | 5.83 ± 2.60 | 6584.5 | 0.88 | 0.98 |
| CTQ physical neglect^a^ | 7.16 ± 2.77 | 7.23 ± 3.10 | 6467.5 | 0.89 | 0.98 |
| CTQ emotional neglect^a^ | 12.53 ± 4.92 | 12.79 ± 4.96 | 6363 | 0.73 | 0.98 |
| CTQ total score^a^ | 41.4 ± 11.36 | 42.59 ± 15.93 | 6660 | 0.80 | 0.98 |
| CQ emotional impact^b^ | 4.05 ± 2.67 | 4.28 ± 2.99 | 5200 | 0.75 | 0.98 |
| CQ attribution of causality^b^ | 5.14 ± 2.41 | 5.36 ± 2.55 | 5074 | 0.54 | 0.98 |
| CQ generalization across time^b^ | 4.41 ± 2.30 | 4.49 ± 2.97 | 5220 | 0.79 | 0.98 |
| CQ generalization across events^b^ | 6.54 ± 3.07 | 5.95 ± 2.90 | 5981 | 0.13 | 0.98 |
| CQ perceived uncontrollability^b^ | 3.91 ± 2.65 | 3.83 ± 2.78 | 5512.5 | 0.68 | 0.98 |
| CQ total score^b^ | 24.15 ± 10.34 | 23.79 ± 11.33 | 5502.5 | 0.70 | 0.98 |

Results are reported as mean ± standard deviation for continuous variables, whereas frequencies are reported for categorical variables. Significant differences are reported in bold. Abbreviations: MDD, Major Depressive Disorder; BD, Bipolar Disorder; HDRS-21, Hamilton Depression Rating Scale – 21 items; CTQ, Childhood Trauma Questionnaire; CQ, Cognition Questionnaire. ^a^subsample of N=229 (108 Cluster 1, 121 Cluster 2); ^b^subsample of N=208 (92 Cluster 1, 116 Cluster 2). *FDR-corrected q<0.05.

### *Table S4. Clinical and demographic comparisons for clusters derived from cortical thickness.*

| **Variable** | **Cluster 1 (N=149)** | **Cluster 2 (N=195)** | **t / U / χ2** | **p** | **q** |
| --- | --- | --- | --- | --- | --- |
| Age | 47.89 ± 10.36 | 48.39 ± 10.42 | 13939 | 0.52 | 0.80 |
| Sex | 93 F; 56 M | 135 F; 60 M | 1.46 | 0.23 | 0.80 |
| Diagnosis | 52 MDD; 97 BD | 83 MDD; 112 BD | 1.77 | 0.18 | 0.80 |
| Scanner | 82 Gyroscan; 67 Igenia | 110 Gyroscan; 85 Igenia | 0.02 | 0.89 | 0.94 |
| No. of episodes | 9.89 ± 11.58 | 9.59 ± 11.04 | 15049 | 0.57 | 0.80 |
| Age at onset | 30.02 ± 10.66 | 30.95 ± 10.76 | 13721.5 | 0.38 | 0.80 |
| Duration of illness (years) | 17.87 ± 10.58 | 17.44 ± 10.98 | 14974 | 0.63 | 0.82 |
| Years of education | 12.12 ± 4.01 | 12.38 ± 3.74 | 13748.5 | 0.42 | 0.80 |
| Pharmacological load | 4.75 ± 2.28 | 4.48 ± 2.16 | 15774 | 0.17 | 0.80 |
| HDRS-21 total score | 22.52 ± 5.83 | 22.39 ± 5.75 | 15153.5 | 0.49 | 0.80 |
| CTQ physical abuse^a^ | 6.10 ± 1.93 | 6.35 ± 4.19 | 6688.5 | 0.68 | 0.83 |
| CTQ emotional abuse^a^ | 8.06 ± 3.50 | 8.91 ± 4.15 | 5871.5 | 0.20 | 0.80 |
| CTQ sexual abuse^a^ | 5.76 ± 2.28 | 5.86 ± 2.58 | 6205 | 0.38 | 0.80 |
| CTQ physical neglect^a^ | 7.11 ± 2.76 | 7.27 ± 3.09 | 6244.5 | 0.58 | 0.80 |
| CTQ emotional neglect^a^ | 12.38 ± 4.97 | 12.90 ± 4.91 | 6175.5 | 0.50 | 0.80 |
| CTQ total score^a^ | 39.99 ± 10.90 | 43.75 ± 15.92 | 5789 | 0.15 | 0.80 |
| CQ emotional impact^b^ | 4.20 ± 2.86 | 4.16 ± 2.86 | 5359.5 | 0.91 | 0.94 |
| CQ attribution of causality^b^ | 5.43 ± 2.31 | 5.14± 2.62 | 5757.5 | 0.30 | 0.80 |
| CQ generalization across time^b^ | 4.59 ± 3.22 | 4.36 ± 2.79 | 5420 | 0.80 | 0.93 |
| CQ generalization across events^b^ | 6.63 ± 2.90 | 5.89 ± 3.01 | 6047 | 0.09 | 0.80 |
| CQ perceived uncontrollability^b^ | 3.87 ± 2.74 | 3.86 ± 2.71 | 5276 | 0.94 | 0.94 |
| CQ total score^b^ | 24.67 ± 10.34 | 23.41 ± 11.28 | 5697 | 0.37 | 0.80 |

Results are reported as mean ± standard deviation for continuous variables, whereas frequencies are reported for categorical variables. Significant differences are reported in bold. Abbreviations: MDD, Major Depressive Disorder; BD, Bipolar Disorder; HDRS-21, Hamilton Depression Rating Scale – 21 items; CTQ, Childhood Trauma Questionnaire; CQ, Cognition Questionnaire. ^a^subsample of N=229 (105 Cluster 1, 124 Cluster 2); ^b^subsample of N=208 (90 Cluster 1, 118 Cluster 2). *FDR-corrected q<0.05.

### *Table S5. Clinical and demographic comparisons for clusters derived from fractional anisotropy.*

| **Variable** | **Cluster 1 (N=121)** | **Cluster 2 (N=154)** | **t / U / χ2** | **p** | **q** |
| --- | --- | --- | --- | --- | --- |
| Age | 48.00 ± 10.00 | 48.00 ± 11.00 | 8918.5 | 0.54 | 0.71 |
| Sex | 78 F; 43 M | 99 F; 55 M | 0 | 1.00 | 1.00 |
| Diagnosis | 35 MDD; 86 BD | 63 MDD; 91 BD | 3.74 | 0.05 | 0.19 |
| Scanner | 81 Gyroscan; 40 Igenia | 88 Gyroscan; 66 Igenia | 2.35 | 0.13 | 0.39 |
| No. of episodes | 11.00 ± 14.00 | 9.50 ± 10.00 | 9524.5 | 0.75 | 0.90 |
| Age at onset | 30.00 ± 12.00 | 30.00 ± 10.00 | 9158 | 0.81 | 0.90 |
| Duration of illness (years) | 17.00 ± 11.00 | 18.00 ± 11.00 | 8926 | 0.55 | 0.71 |
| Years of education | 13.00 ± 3.90 | 12.00 ± 3.80 | 10594 | **0.04** | 0.19 |
| Pharmacological load | 4.40 ± 2.20 | 4.50 ± 2.20 | 9166.5 | 0.82 | 0.90 |
| HDRS-21 total score | 23.00 ± 5.90 | 22.00 ± 5.60 | 9940.5 | 0.34 | 0.59 |
| CTQ physical abuse^a^ | 5.80 ± 2.10 | 6.60 ± 4.50 | 3161.5 | **0.01** | 0.19 |
| CTQ emotional abuse^a^ | 8.10 ± 3.90 | 8.70 ± 4.00 | 3473.5 | 0.20 | 0.49 |
| CTQ sexual abuse^a^ | 5.80 ± 2.70 | 5.90 ± 2.60 | 3686.5 | 0.35 | 0.59 |
| CTQ physical neglect^a^ | 6.70 ± 2.60 | 7.30 ± 3.00 | 3480.5 | 0.20 | 0.49 |
| CTQ emotional neglect^a^ | 12.00 ± 5.30 | 13.00 ± 4.80 | 3610 | 0.39 | 0.61 |
| CTQ total score^a^ | 39.00 ± 13.00 | 42.00 ± 12.00 | 3232.5 | 0.05 | 0.19 |
| CQ emotional impact^b^ | 3.60 ± 3.10 | 4.40 ± 2.80 | 2670 | **0.03** | 0.19 |
| CQ attribution of causality^b^ | 4.80 ± 2.40 | 5.60 ± 2.60 | 2729 | 0.05 | 0.19 |
| CQ generalization across time^b^ | 4.20 ± 2.80 | 4.80 ± 3.10 | 3000.5 | 0.27 | 0.56 |
| CQ generalization across events^b^ | 6.10 ± 3.10 | 6.40 ± 2.90 | 3105 | 0.46 | 0.67 |
| CQ perceived uncontrollability^b^ | 3.90 ± 2.70 | 3.80 ± 2.80 | 3377 | 0.88 | 0.92 |
| CQ total score^b^ | 23.00 ± 11.00 | 25.00 ± 11.00 | 3003.5 | 0.28 | 0.56 |

Results are reported as mean ± standard deviation for continuous variables, whereas frequencies are reported for categorical variables. Significant differences are reported in bold. Abbreviations: MDD, Major Depressive Disorder; BD, Bipolar Disorder; HDRS-21, Hamilton Depression Rating Scale – 21 items; CTQ, Childhood Trauma Questionnaire; CQ, Cognition Questionnaire. ^a^subsample of N=180 (73 Cluster 1, 107 Cluster 2); ^b^subsample of N=164 (74 Cluster 1, 90 Cluster 2). *FDR-corrected q<0.05.

### *Table S6. Clinical and demographic comparisons for clusters derived from axial diffusivity.*

| **Variable** | **Cluster 1 (N=120)** | **Cluster 2 (N=155)** | **t / U / χ2** | **p** | **q** |
| --- | --- | --- | --- | --- | --- |
| Age | 48.51 ± 10.62 | 47.45 ± 10.41 | 9938.5 | 0.33 | 0.88 |
| Sex | 87 F; 33 M | 90 F; 65 M | 5.53 | **0.02** | 0.21 |
| Diagnosis | 34 MDD; 86 BD | 64 MDD; 91 BD | 4.4 | **0.04** | 0.26 |
| Scanner | 77 Gyroscan; 43 Igenia | 92 Gyroscan; 63 Igenia | 0.47 | 0.49 | 0.90 |
| No. of episodes | 10.71 ± 12.22 | 9.619 ± 11.35 | 10271 | 0.14 | 0.77 |
| Age at onset | 31.12 ± 11.19 | 29.66 ± 10.69 | 9908.5 | 0.35 | 0.88 |
| Duration of illness (years) | 17.39 ± 10.72 | 17.78 ± 10.95 | 9102.5 | 0.76 | 0.94 |
| Years of education | 11.57 ± 3.917 | 12.77 ± 3.713 | 7753.5 | **0.01** | 0.21 |
| Pharmacological load | 4.583 ± 2.371 | 4.387 ± 2.027 | 9579 | 0.67 | 0.92 |
| HDRS-21 total score | 22.61 ± 5.607 | 22.63 ± 5.882 | 9286.5 | 0.98 | 0.99 |
| CTQ physical abuse^a^ | 6.46 ± 4.92 | 6.23 ± 2.64 | 3762.5 | 0.67 | 0.92 |
| CTQ emotional abuse^a^ | 8.40 ± 4.09 | 8.55 ± 3.83 | 3677 | 0.53 | 0.90 |
| CTQ sexual abuse^a^ | 5.78 ± 2.30 | 5.91 ± 2.82 | 3945 | 0.81 | 0.94 |
| CTQ physical neglect^a^ | 6.99 ± 2.61 | 7.07 ± 3.02 | 3984 | 0.77 | 0.94 |
| CTQ emotional neglect^a^ | 12.19 ± 4.58 | 13.01 ± 5.31 | 3577.5 | 0.36 | 0.88 |
| CTQ total score^a^ | 39.35 ± 11.03 | 41.19 ± 13.29 | 3614.5 | 0.43 | 0.90 |
| CQ emotional impact^b^ | 3.79 ± 2.86 | 4.24 ± 2.99 | 2893.5 | 0.33 | 0.88 |
| CQ attribution of causality^b^ | 5.46 ± 2.75 | 5.14 ± 2.45 | 3390 | 0.48 | 0.90 |
| CQ generalization across time^b^ | 4.52 ± 2.98 | 4.57 ± 2.99 | 3184.5 | 0.99 | 0.99 |
| CQ generalization across events^b^ | 5.87 ± 2.90 | 6.55 ± 3.01 | 2811.5 | 0.21 | 0.88 |
| CQ perceived uncontrollability^b^ | 3.73 ± 2.51 | 3.89 ± 2.92 | 3156.5 | 0.93 | 0.99 |
| CQ total score^b^ | 23.24 ± 11.00 | 24.30 ± 11.25 | 3030 | 0.61 | 0.92 |

Results are reported as mean ± standard deviation for continuous variables, whereas frequencies are reported for categorical variables. Significant differences are reported in bold. Abbreviations: MDD, Major Depressive Disorder; BD, Bipolar Disorder; HDRS-21, Hamilton Depression Rating Scale – 21 items; CTQ, Childhood Trauma Questionnaire; CQ, Cognition Questionnaire. ^a^subsample of N=180 (72 Cluster 1,108 Cluster 2); ^b^subsample of N=164 (63 Cluster 1, 101 Cluster 2). *FDR-corrected q<0.05.

### *Table S7. Clinical and demographic comparisons for clusters derived from radial diffusivity.*

| **Variable** | **Cluster 1 (N=137)** | **Cluster 2 (N=138)** | **t / U / χ2** | **p** | **q** |
| --- | --- | --- | --- | --- | --- |
| Age | 46.77 ± 10.80 | 49.04 ± 10.10 | 8338 | 0.09 | 0.40 |
| Sex | 92 F; 45 M | 85 F; 53 M | 0.7 | 0.40 | 0.89 |
| Diagnosis | 50 MDD; 87 BD | 48 MDD; 90 BD | 0.03 | 0.86 | 0.90 |
| Scanner | 83 Gyroscan; 54 Igenia | 86 Gyroscan; 52 Igenia | 0.03 | 0.86 | 0.90 |
| No. of episodes | 9.69 ± 11.59 | 10.49 ± 11.89 | 9050.5 | 0.54 | 0.90 |
| Age at onset | 28.89 ± 10.68 | 31.7 ± 11.01 | 8025 | **0.03** | 0.33 |
| Duration of illness (years) | 17.88 ± 10.89 | 17.34 ± 10.81 | 9719 | 0.69 | 0.90 |
| Years of education | 12.90 ± 3.65 | 11.60 ± 3.94 | 11179.5 | **0.01** | 0.13 |
| Pharmacological load | 4.71 ± 2.20 | 4.24 ± 2.15 | 10604.5 | 0.08 | 0.40 |
| HDRS-21 total score | 22.66 ± 5.47 | 22.58 ± 6.04 | 9650.5 | 0.76 | 0.90 |
| CTQ physical abuse^a^ | 5.98 ± 2.61 | 6.67 ± 4.55 | 3460.5 | 0.05 | 0.38 |
| CTQ emotional abuse^a^ | 8.62 ± 4.02 | 8.36 ± 3.85 | 4251 | 0.56 | 0.90 |
| CTQ sexual abuse^a^ | 5.96 ± 2.96 | 5.76 ± 2.23 | 4021 | 0.90 | 0.90 |
| CTQ physical neglect^a^ | 7.16 ± 3.16 | 6.92 ± 2.53 | 4111 | 0.86 | 0.90 |
| CTQ emotional neglect^a^ | 12.88 ± 5.39 | 12.49 ± 4.67 | 4215 | 0.64 | 0.90 |
| CTQ total score^a^ | 41.10 ± 14.01 | 39.81 ± 10.67 | 4101 | 0.89 | 0.90 |
| CQ emotional impact^b^ | 4.26 ± 3.17 | 3.86 ± 2.68 | 3502 | 0.63 | 0.90 |
| CQ attribution of causality^b^ | 5.20 ± 2.40 | 5.33 ± 2.74 | 3300.5 | 0.85 | 0.90 |
| CQ generalization across time^b^ | 4.79 ± 3.02 | 4.30 ± 2.93 | 3672.5 | 0.30 | 0.82 |
| CQ generalization across events^b^ | 6.58 ± 3.18 | 5.99 ± 2.73 | 3635.5 | 0.36 | 0.88 |
| CQ perceived uncontrollability^b^ | 4.18 ± 2.84 | 3.46 ± 2.64 | 3841 | 0.11 | 0.40 |
| CQ total score^b^ | 25.00 ± 11.48 | 22.70 ± 10.69 | 3743 | 0.20 | 0.64 |

Results are reported as mean ± standard deviation for continuous variables, whereas frequencies are reported for categorical variables. Significant differences are reported in bold. Abbreviations: MDD, Major Depressive Disorder; BD, Bipolar Disorder; HDRS-21, Hamilton Depression Rating Scale – 21 items; CTQ, Childhood Trauma Questionnaire; CQ, Cognition Questionnaire. ^a^subsample of N=180 (90 Cluster 1, 90 Cluster 2); ^b^subsample of N=164 (85 Cluster 1, 79 Cluster 2). *FDR-corrected q<0.05.

### *Table S8. Clinical and demographic comparisons for clusters derived from mean diffusivity.*

| **Variable** | **Cluster 1 (N=166)** | **Cluster 2 (N=109)** | **t / U / χ2** | **p** | **q** |
| --- | --- | --- | --- | --- | --- |
| Age | 48.2 ± 10.64 | 47.47 ± 10.32 | 9502.5 | 0.48 | 0.75 |
| Sex | 112 F; 54 M | 65 F; 44 M | 1.44 | 0.23 | 0.70 |
| Diagnosis | 66 MDD; 100 BD | 32 MDD; 77 BD | 2.67 | 0.10 | 0.55 |
| Scanner | 99 Gyroscan; 67 Igenia | 70 Gyroscan; 39 Igenia | 0.41 | 0.52 | 0.77 |
| No. of episodes | 9.337 ± 11.16 | 11.25 ± 12.51 | 7874 | 0.07 | 0.50 |
| Age at onset | 30.18 ± 11.27 | 30.48 ± 10.4 | 8799 | 0.70 | 0.79 |
| Duration of illness (years) | 18.02 ± 10.97 | 16.99 ± 10.63 | 9505.5 | 0.48 | 0.75 |
| Years of education | 12.82 ± 3.683+ | 11.38 ± 3.93 | 10862 | **<0.01** | 0.07 |
| Pharmacological load | 4.57 ± 2.17 | 4.33 ± 2.20 | 9621.5 | 0.37 | 0.70 |
| HDRS-21 total score | 22.42 ± 5.76 | 22.92 ± 5.75 | 8884.5 | 0.80 | 0.84 |
| CTQ physical abuse^a^ | 5.96 ± 2.46 | 6.90 ± 5.10 | 3171.5 | **0.03** | 0.28 |
| CTQ emotional abuse^a^ | 8.63 ± 4.16 | 8.26 ± 3.54 | 3950 | 0.72 | 0.79 |
| CTQ sexual abuse^a^ | 6.06 ± 3.03 | 5.52 ± 1.73 | 3932.5 | 0.65 | 0.79 |
| CTQ physical neglect^a^ | 7.23 ± 3.12 | 6.74 ± 2.37 | 4113.5 | 0.38 | 0.70 |
| CTQ emotional neglect^a^ | 12.97 ± 5.32 | 12.22 ± 4.53 | 4134.5 | 0.37 | 0.70 |
| CTQ total score^a^ | 41.27 ± 13.81 | 39.14 ± 9.80 | 3993.5 | 0.63 | 0.79 |
| CQ emotional impact^b^ | 4.03 ± 3.05 | 4.14 ± 2.75 | 2870 | 0.59 | 0.79 |
| CQ attribution of causality^b^ | 5.02 ± 2.42 | 5.73 ± 2.79 | 2584.5 | 0.13 | 0.55 |
| CQ generalization across time^b^ | 4.40 ± 2.93 | 4.86 ± 3.07 | 2748 | 0.34 | 0.70 |
| CQ generalization across events^b^ | 6.16 ± 3.10 | 6.55 ± 2.73 | 2683 | 0.23 | 0.70 |
| CQ perceived uncontrollability^b^ | 3.88 ± 2.81 | 3.73 ± 2.69 | 3080.5 | 0.84 | 0.84 |
| CQ total score^b^ | 23.35 ± 11.20 | 24.93 ± 11.04 | 2756.5 | 0.35 | 0.70 |

Results are reported as mean ± standard deviation for continuous variables, whereas frequencies are reported for categorical variables. Significant differences are reported in bold. Abbreviations: MDD, Major Depressive Disorder; BD, Bipolar Disorder; HDRS-21, Hamilton Depression Rating Scale – 21 items; CTQ, Childhood Trauma Questionnaire; CQ, Cognition Questionnaire. ^a^subsample of N=180 (111 Cluster 1, 69 Cluster 2); ^b^subsample of N=164 (108 Cluster 1, 56 Cluster 2). *FDR-corrected q<0.05.

### *Table S9. Clinical and demographic comparisons for the multimodal-derived clusters.*

| **Variable** | **Cluster 1 (N=59)** | **Cluster 2 (N=57)** | **t / U / χ2** | **p** | **q** |
| --- | --- | --- | --- | --- | --- |
| Age | 47.98 ± 12.29 | 46.35 ± 10.07 | 1871.50 | 0.30 | 0.66 |
| Sex | 38 F; 21 M | 36 F; 21 M | 0 | 1.00 | 1.00 |
| Diagnosis | 26 MDD; 33 BD | 30 MDD; 27 BD | 0.54 | 0.46 | 0.76 |
| Scanner | 27 Gyroscan; 32 Igenia | 22 Gyroscan; 35 Igenia | 0.35 | 0.55 | 0.76 |
| No. of episodes | 10.66 ± 12.51 | 8.72 ± 11.00 | 1874.50 | 0.28 | 0.66 |
| Age at onset | 30.17 ± 11.57 | 28.42 ± 10.26 | 1799.00 | 0.52 | 0.76 |
| Duration of illness (years) | 17.81 ± 11.50 | 17.93 ± 10.75 | 1683.50 | 0.99 | 1.00 |
| Years of education | 13.02 ± 3.51 | 12.23 ± 3.47 | 1882.00 | 0.24 | 0.66 |
| Pharmacological load | 4.85 ± 2.15 | 4.72 ± 2.14 | 1771.00 | 0.62 | 0.76 |
| HDRS-21 total score | 21.80 ± 5.35 | 21.44 ± 6.31 | 0.33 | 0.74 | 0.81 |
| CTQ physical abuse^a^ | 6.34 ± 3.39 | 6.10 ± 2.03 | 1049.00 | 0.66 | 0.76 |
| CTQ emotional abuse^a^ | 9.23 ± 4.49 | 8.66 ± 4.02 | 1177.50 | 0.55 | 0.76 |
| CTQ sexual abuse^a^ | 6.50 ± 3.74 | 5.72 ± 2.44 | 1205.50 | 0.26 | 0.66 |
| CTQ physical neglect^a^ | 7.73 ± 3.91 | 6.76 ± 2.48 | 1217.00 | 0.35 | 0.70 |
| CTQ emotional neglect^a^ | 13.23 ± 5.69 | 12.26 ± 4.74 | 1207.00 | 0.42 | 0.76 |
| CTQ total score^a^ | 43.34 ± 17.26 | 39.50 ± 10.36 | 1166.50 | 0.62 | 0.76 |
| CQ emotional impact^b^ | 4.85 ± 3.30 | 3.69 ± 2.26 | 915.50 | 0.18 | 0.66 |
| CQ attribution of causality^b^ | 5.65 ± 2.44 | 5.18 ± 2.10 | 884.50 | 0.30 | 0.66 |
| CQ generalization across time^b^ | 5.13 ± 3.12 | 4.18 ± 2.78 | 935.00 | 0.13 | 0.66 |
| CQ generalization across events^b^ | 7.03 ± 2.70 | 5.95 ± 2.82 | 1.73 | 0.09 | 0.66 |
| CQ perceived uncontrollability^b^ | 4.48 ± 3.04 | 3.00 ± 2.44 | 998.50 | **0.03** | 0.55 |
| CQ total score^b^ | 27.12 ± 11.75 | 21.90 ± 9.12 | 981.00 | 0.05 | 0.55 |

Results are reported as mean ± standard deviation for continuous variables, whereas frequencies are reported for categorical variables. Significant differences are reported in bold. Abbreviations: MDD, Major Depressive Disorder; BD, Bipolar Disorder; HDRS-21, Hamilton Depression Rating Scale – 21 items; CTQ, Childhood Trauma Questionnaire; CQ, Cognition Questionnaire. ^a^subsample of N=94 (44 Cluster 1, 50 Cluster 2); ^b^subsample of N=79 (40 Cluster 1, 39 Cluster 2). *FDR-corrected q<0.05.

### *Table S10. Clinical and demographic comparisons with healthy controls for the FC-based clusters*

| **Variable** | **HC (N=138)** | **Cluster 1 (N=50)** | **Cluster 2 (N=96)** | **F / χ2** | **p** | **pFDR** | **post-hoc** |
| --- | --- | --- | --- | --- | --- | --- | --- |
| Age | 30.75±10.6 | 47.5±10.11 | 47.67±11.57 | 85.49 | <0.001 | **<0.001*** | 1>HC 2>HC |
| Sex | 77 F; 61 M | 35 F; 15 M | 64 F; 32 M | 4.50 | 0.11 | 0.18 |  |
| Scanner | 0 Gyroscan; 138 Igenia | 16 Gyroscan; 34 Igenia | 34 Gyroscan; 62 Igenia | 57.62 | <0.001 | **<0.001*** |  |
| No. of episodes | / | 7.98±10.21 | 9.47±11.49 | 0.59 | 0.44 | 0.51 |  |
| Age at onset | / | 30.36±10.72 | 29.2±10.43 | 0.40 | 0.53 | 0.56 |  |
| Duration of illness (years) | / | 17.14±10.71 | 18.47±11.09 | 0.48 | 0.49 | 0.54 |  |
| Years of education | 16.51±3.04 | 12.98±3.24 | 12.74±3.69 | 43.95 | <0.001 | **<0.001*** | HC>1 HC>2 |
| Pharmacological load | / | 4.54±2.05 | 4.89±2.26 | 0.82 | 0.37 | 0.46 |  |
| HDRS-21 total score | / | 21.96±5.98 | 21.49±6.17 | 0.20 | 0.66 | 0.66 |  |
| CTQ physical abuse^a^ | 5.62±1.58 | 5.82±1.99 | 6.23±2.68 | 1.86 | 0.16 | 0.24 |  |
| CTQ emotional abuse^a^ | 8.04±3.59 | 8.33±4.25 | 9.05±3.92 | 1.57 | 0.21 | 0.29 |  |
| CTQ sexual abuse^a^ | 5.06±0.78 | 5.44±1.96 | 6.22±3.15 | 6.50 | <0.001 | **<0.001*** | 2>HC |
| CTQ physical neglect^a^ | 6.58±2.74 | 7.18±2.92 | 7.3±3.29 | 1.44 | 0.24 | 0.32 |  |
| CTQ emotional neglect^a^ | 11.1±4.56 | 12.15±4.53 | 12.68±5.19 | 2.50 | 0.08 | 0.14 |  |
| CTQ total score^a^ | 36.77±10.07 | 41.31±13.25 | 43.17±15.54 | 5.74 | <0.001 | **<0.001*** | 2>HC |
| CQ emotional impact^b^ | 3.04±3.17 | 4.51±2.7 | 4.28±2.78 | 3.57 | 0.03 | 0.06 |  |
| CQ attribution of causality^b^ | 3.5±2.2 | 5.06±2.22 | 5.59±2.23 | 13.24 | <0.001 | **<0.001*** | 1>HC 2>HC |
| CQ generalization across time^b^ | 2.74±1.83 | 3.89±2.89 | 4.72±2.86 | 8.58 | <0.001 | **<0.001*** | 2>HC |
| CQ generalization across events^b^ | 3.76±2.51 | 5.37±2.92 | 6.59±2.76 | 15.76 | <0.001 | **<0.001*** | 1>HC 2>HC |
| CQ perceived uncontrollability^b^ | 2.68±2.05 | 2.97±2.7 | 4.23±2.69 | 6.37 | <0.001 | **<0.001*** | 2>HC |
| CQ total score^b^ | 14.26±7.71 | 21.69±10.7 | 25.42±10.2 | 19.78 | <0.001 | **<0.001*** | 1>HC 2>HC |

Results are reported as mean ± standard deviation for continuous variables, whereas frequencies are reported for categorical variables. Significant differences are reported in bold. Abbreviations: HC, Healthy Controls; MDD, Major Depressive Disorder; BD, Bipolar Disorder; HDRS-21, Hamilton Depression Rating Scale – 21 items; CTQ, Childhood Trauma Questionnaire; CQ, Cognition Questionnaire. ^a^subsample of N=219 (39 Cluster 1, 82 Cluster 2, 98 HC); ^b^subsample of N=154 (35 Cluster 1, 69 Cluster 2, 50 HC). *FDR-corrected q<0.05.

### *Table S11. Results of grouped PLSC between the FC-based clusters*

| **LV** | **Explained covariance** | **p** | **pFDR** |
| --- | --- | --- | --- |
| LV1 | 0.40 | 0.001 | 0.01* |
| LV2 | 0.26 | 0.841 | 0.998 |
| LV3 | 0.10 | 0.581 | 0.998 |
| LV4 | 0.07 | 0.339 | 0.998 |
| LV5 | 0.05 | 0.623 | 0.998 |
| LV6 | 0.05 | 0.729 | 0.998 |
| LV7 | 0.03 | 0.837 | 0.998 |
| LV8 | 0.02 | 0.985 | 0.998 |
| LV9 | 0.02 | 0.998 | 0.998 |
| LV10 | 0.01 | 0.974 | 0.998 |

Abbreviations: LV, Latent variable. *FDR-corrected q<0.05.

### *Table S12. Results of grouped PLSC between the FC-based clusters and healthy controls*

| **LV** | **Explained covariance** | **p** | **pFDR** |
| --- | --- | --- | --- |
| LV1 | 0.43 | 0.001 | 0.015* |
| LV2 | 0.16 | 0.524 | 1 |
| LV3 | 0.10 | 0.746 | 1 |
| LV4 | 0.07 | 0.494 | 1 |
| LV5 | 0.05 | 0.656 | 1 |
| LV6 | 0.04 | 0.626 | 1 |
| LV7 | 0.04 | 0.959 | 1 |
| LV8 | 0.03 | 0.909 | 1 |
| LV9 | 0.02 | 0.978 | 1 |
| LV10 | 0.02 | 0.969 | 1 |
| LV11 | 0.01 | 0.987 | 1 |
| LV12 | 0.01 | 0.99 | 1 |
| LV13 | 0.01 | 0.999 | 1 |
| LV14 | 0.01 | 1 | 1 |
| LV15 | 0.00 | 0.998 | 1 |

Abbreviations: LV, Latent variable. *FDR-corrected q<0.05.

### *Table S13. Cluster differences in regional positive and negative FC strength compared to healthy controls.*

|  |  | **HC vs Cluster 1 vs Cluster 2** | | | | **HC vs Cluster 1** | | | | **HC vs Cluster 2** | | | | **Cluster 1 vs Cluster 2** | | | |
| --- | --- | --- | --- | --- | --- | --- | --- | --- | --- | --- | --- | --- | --- | --- | --- | --- | --- |
| **Brainettome label** | **Measure** | **F** | **p** | **eta2** | **q** | **t** | **p** | **d** | **q** | **t** | **p** | **d** | **q** | **t** | **p** | **d** | **q** |
| SFG medial frontal area 8 L | Spos | 220.33 | <0.001 | 0.61 | <0.001 | -13.79 | <0.001 | -3.22 | <0.001 | -11.35 | <0.001 | -1.55 | <0.001 | 8.75 | <0.001 | 1.86 | <0.001 |
| SFG medial frontal area 8 R | Spos | 273.33 | <0.001 | 0.66 | <0.001 | -15.67 | <0.001 | -3.56 | <0.001 | -12.28 | <0.001 | -1.65 | <0.001 | 10.12 | <0.001 | 2.15 | <0.001 |
| SFG dorsolateral area 6 L | Spos | 209.19 | <0.001 | 0.60 | <0.001 | -13.54 | <0.001 | -3.20 | <0.001 | -11.23 | <0.001 | -1.57 | <0.001 | 8.27 | <0.001 | 1.73 | <0.001 |
| SFG medial area 6 L | Spos | 294.78 | <0.001 | 0.68 | <0.001 | -16.62 | <0.001 | -3.79 | <0.001 | -10.78 | <0.001 | -1.48 | <0.001 | 11.34 | <0.001 | 2.36 | <0.001 |
| SFG medial area 6 R | Spos | 326.37 | <0.001 | 0.70 | <0.001 | -17.95 | <0.001 | -4.07 | <0.001 | -10.28 | <0.001 | -1.42 | <0.001 | 12.43 | <0.001 | 2.54 | <0.001 |
| SFG medial area 9 L | Spos | 240.67 | <0.001 | 0.63 | <0.001 | -14.68 | <0.001 | -3.35 | <0.001 | -11.82 | <0.001 | -1.60 | <0.001 | 9.29 | <0.001 | 1.96 | <0.001 |
| SFG medial area 9 R | Spos | 254.05 | <0.001 | 0.64 | <0.001 | -15.22 | <0.001 | -3.48 | <0.001 | -12.65 | <0.001 | -1.73 | <0.001 | 9.20 | <0.001 | 1.92 | <0.001 |
| IFG dorsolateral area 44 R | Spos | 248.61 | <0.001 | 0.64 | <0.001 | -14.64 | <0.001 | -3.49 | <0.001 | -14.16 | <0.001 | -2.01 | <0.001 | 7.87 | <0.001 | 1.63 | <0.001 |
| IFG opercular area 44 L | Spos | 161.36 | <0.001 | 0.53 | <0.001 | -11.55 | <0.001 | -2.77 | <0.001 | -11.28 | <0.001 | -1.59 | <0.001 | 6.52 | <0.001 | 1.37 | <0.001 |
| IFG opercular area 44 R | Spos | 179.90 | <0.001 | 0.56 | <0.001 | -12.75 | <0.001 | -2.98 | <0.001 | -10.87 | <0.001 | -1.52 | <0.001 | 7.43 | <0.001 | 1.53 | <0.001 |
| PreCG area 4 upperimb 4 L | Spos | 272.37 | <0.001 | 0.66 | <0.001 | -16.69 | <0.001 | -3.74 | <0.001 | -10.80 | <0.001 | -1.50 | <0.001 | 10.76 | <0.001 | 2.18 | <0.001 |
| ParaCL area 1/2/3owerimb R | Spos | 294.10 | <0.001 | 0.68 | <0.001 | -17.09 | <0.001 | -3.86 | <0.001 | -10.27 | <0.001 | -1.42 | <0.001 | 11.60 | <0.001 | 2.37 | <0.001 |
| ParaCL area 4owerimb L | Spos | 334.23 | <0.001 | 0.70 | <0.001 | -18.36 | <0.001 | -4.05 | <0.001 | -11.03 | <0.001 | -1.50 | <0.001 | 12.59 | <0.001 | 2.59 | <0.001 |
| ParaCL area 4owerimb R | Spos | 346.10 | <0.001 | 0.71 | <0.001 | -19.27 | <0.001 | -4.18 | <0.001 | -10.53 | <0.001 | -1.44 | <0.001 | 13.32 | <0.001 | 2.69 | <0.001 |
| FusGy medioventral area 37 L | Spos | 263.30 | <0.001 | 0.65 | <0.001 | -17.05 | <0.001 | -3.58 | <0.001 | -11.88 | <0.001 | -1.60 | <0.001 | 10.42 | <0.001 | 2.10 | <0.001 |
| FusGy medioventral area 37 R | Spos | 241.01 | <0.001 | 0.63 | <0.001 | -15.58 | <0.001 | -3.36 | <0.001 | -12.08 | <0.001 | -1.61 | <0.001 | 9.47 | <0.001 | 1.96 | <0.001 |
| ParHipGyateral post PH area L | Spos | 144.80 | <0.001 | 0.51 | <0.001 | -12.89 | <0.001 | -2.80 | <0.001 | -7.82 | <0.001 | -1.09 | <0.001 | 7.99 | <0.001 | 1.56 | <0.001 |
| ParHipGyateral post PH area R | Spos | 118.49 | <0.001 | 0.46 | <0.001 | -11.31 | <0.001 | -2.49 | <0.001 | -9.79 | <0.001 | -1.38 | <0.001 | 5.27 | <0.001 | 1.02 | <0.001 |
| ParHipGy Tharea medialPHG L | Spos | 80.05 | <0.001 | 0.36 | <0.001 | -8.77 | <0.001 | -1.99 | <0.001 | -7.61 | <0.001 | -1.06 | <0.001 | 4.78 | <0.001 | 0.97 | <0.001 |
| ParHipGy Tharea medialPHG R | Spos | 92.19 | <0.001 | 0.40 | <0.001 | -8.24 | <0.001 | -1.99 | <0.001 | -8.62 | <0.001 | -1.16 | <0.001 | 5.20 | <0.001 | 1.15 | <0.001 |
| PostSupTempSulc caudopost sup temp sulc L | Spos | 219.50 | <0.001 | 0.61 | <0.001 | -14.76 | <0.001 | -3.21 | <0.001 | -13.34 | <0.001 | -1.81 | <0.001 | 7.73 | <0.001 | 1.57 | <0.001 |
| SPLateral area 5 L | Spos | 252.01 | <0.001 | 0.64 | <0.001 | -15.72 | <0.001 | -3.52 | <0.001 | -12.65 | <0.001 | -1.74 | <0.001 | 9.14 | <0.001 | 1.87 | <0.001 |
| SPLateral area 5 R | Spos | 285.46 | <0.001 | 0.67 | <0.001 | -16.04 | <0.001 | -3.68 | <0.001 | -13.19 | <0.001 | -1.80 | <0.001 | 9.88 | <0.001 | 2.07 | <0.001 |
| SPL intrapariet area 7 R | Spos | 265.63 | <0.001 | 0.65 | <0.001 | -15.51 | <0.001 | -3.59 | <0.001 | -13.24 | <0.001 | -1.84 | <0.001 | 9.13 | <0.001 | 1.90 | <0.001 |
| PreCun medial area 7 L | Spos | 294.54 | <0.001 | 0.68 | <0.001 | -18.64 | <0.001 | -3.89 | <0.001 | -10.63 | <0.001 | -1.44 | <0.001 | 12.17 | <0.001 | 2.41 | <0.001 |
| PreCun medial area 7 R | Spos | 290.28 | <0.001 | 0.67 | <0.001 | -17.59 | <0.001 | -3.84 | <0.001 | -11.38 | <0.001 | -1.56 | <0.001 | 11.21 | <0.001 | 2.26 | <0.001 |
| PreCun medial area 5 L | Spos | 296.57 | <0.001 | 0.68 | <0.001 | -19.50 | <0.001 | -4.01 | <0.001 | -9.05 | <0.001 | -1.24 | <0.001 | 13.26 | <0.001 | 2.55 | <0.001 |
| PreCun medial area 5 R | Spos | 368.11 | <0.001 | 0.72 | <0.001 | -21.81 | <0.001 | -4.55 | <0.001 | -10.42 | <0.001 | -1.45 | <0.001 | 14.38 | <0.001 | 2.74 | <0.001 |
| PreCun dorsomed parietocc sulcus L | Spos | 232.76 | <0.001 | 0.62 | <0.001 | -16.67 | <0.001 | -3.50 | <0.001 | -10.43 | <0.001 | -1.43 | <0.001 | 10.12 | <0.001 | 1.98 | <0.001 |
| PreCun dorsomed parietocc sulcus R | Spos | 232.96 | <0.001 | 0.62 | <0.001 | -15.81 | <0.001 | -3.42 | <0.001 | -11.27 | <0.001 | -1.54 | <0.001 | 9.48 | <0.001 | 1.90 | <0.001 |
| PreCun area 31 L | Spos | 196.16 | <0.001 | 0.58 | <0.001 | -14.30 | <0.001 | -3.18 | <0.001 | -11.20 | <0.001 | -1.56 | <0.001 | 7.93 | <0.001 | 1.58 | <0.001 |
| PreCun area 31 R | Spos | 242.88 | <0.001 | 0.63 | <0.001 | -16.90 | <0.001 | -3.55 | <0.001 | -11.62 | <0.001 | -1.59 | <0.001 | 9.70 | <0.001 | 1.90 | <0.001 |
| InsL hypergranular insular L | Spos | 134.25 | <0.001 | 0.49 | <0.001 | -11.99 | <0.001 | -2.67 | <0.001 | -8.55 | <0.001 | -1.20 | <0.001 | 6.96 | <0.001 | 1.38 | <0.001 |
| InsL hypergranular insular R | Spos | 130.89 | <0.001 | 0.48 | <0.001 | -11.25 | <0.001 | -2.54 | <0.001 | -7.81 | <0.001 | -1.07 | <0.001 | 7.28 | <0.001 | 1.50 | <0.001 |
| InsL ventral agranular insular L | Spos | 85.75 | <0.001 | 0.38 | <0.001 | -9.76 | <0.001 | -2.14 | <0.001 | -7.35 | <0.001 | -1.03 | <0.001 | 5.23 | <0.001 | 1.02 | <0.001 |
| InsL ventral agranular insular R | Spos | 58.00 | <0.001 | 0.29 | <0.001 | -7.00 | <0.001 | -1.66 | <0.001 | -6.73 | <0.001 | -0.94 | <0.001 | 3.94 | <0.001 | 0.82 | <0.001 |
| InsL dorsolat agranular insular L | Spos | 162.62 | <0.001 | 0.54 | <0.001 | -11.39 | <0.001 | -2.71 | <0.001 | -11.48 | <0.001 | -1.58 | <0.001 | 6.69 | <0.001 | 1.44 | <0.001 |
| InsL dorsolat agranular insular R | Spos | 168.68 | <0.001 | 0.55 | <0.001 | -12.68 | <0.001 | -2.90 | <0.001 | -9.16 | <0.001 | -1.27 | <0.001 | 8.03 | <0.001 | 1.65 | <0.001 |
| InsL ventral granular insular L | Spos | 123.38 | <0.001 | 0.47 | <0.001 | -10.39 | <0.001 | -2.43 | <0.001 | -8.50 | <0.001 | -1.17 | <0.001 | 6.50 | <0.001 | 1.37 | <0.001 |
| InsL ventral granular insular R | Spos | 147.91 | <0.001 | 0.51 | <0.001 | -11.41 | <0.001 | -2.68 | <0.001 | -9.26 | <0.001 | -1.29 | <0.001 | 7.09 | <0.001 | 1.48 | <0.001 |
| InsL dorsolat granular insular L | Spos | 129.79 | <0.001 | 0.48 | <0.001 | -10.85 | <0.001 | -2.52 | <0.001 | -8.94 | <0.001 | -1.25 | <0.001 | 6.51 | <0.001 | 1.35 | <0.001 |
| InsL dorsolat granular insular R | Spos | 137.30 | <0.001 | 0.49 | <0.001 | -11.28 | <0.001 | -2.61 | <0.001 | -9.13 | <0.001 | -1.27 | <0.001 | 6.74 | <0.001 | 1.39 | <0.001 |
| InsL dorsal dysgranular insular L | Spos | 165.50 | <0.001 | 0.54 | <0.001 | -12.14 | <0.001 | -2.83 | <0.001 | -9.52 | <0.001 | -1.31 | <0.001 | 7.69 | <0.001 | 1.61 | <0.001 |
| InsL dorsal dysgranular insular R | Spos | 169.69 | <0.001 | 0.55 | <0.001 | -12.87 | <0.001 | -2.96 | <0.001 | -9.64 | <0.001 | -1.35 | <0.001 | 7.73 | <0.001 | 1.57 | <0.001 |
| CingGy dorsal area 23 L | Spos | 229.90 | <0.001 | 0.62 | <0.001 | -16.00 | <0.001 | -3.38 | <0.001 | -10.09 | <0.001 | -1.36 | <0.001 | 10.28 | <0.001 | 2.07 | <0.001 |
| CingGy dorsal area 23 R | Spos | 213.28 | <0.001 | 0.60 | <0.001 | -14.99 | <0.001 | -3.23 | <0.001 | -9.88 | <0.001 | -1.33 | <0.001 | 9.68 | <0.001 | 1.98 | <0.001 |
| CingGyostroventral area 24 L | Spos | 117.06 | <0.001 | 0.45 | <0.001 | -10.33 | <0.001 | -2.44 | <0.001 | -8.10 | <0.001 | -1.14 | <0.001 | 6.29 | <0.001 | 1.29 | <0.001 |
| CingGy pregenual area 32 L | Spos | 159.63 | <0.001 | 0.53 | <0.001 | -12.09 | <0.001 | -2.79 | <0.001 | -8.52 | <0.001 | -1.17 | <0.001 | 7.99 | <0.001 | 1.67 | <0.001 |
| CingGy pregenual area 32 R | Spos | 150.77 | <0.001 | 0.52 | <0.001 | -11.08 | <0.001 | -2.62 | <0.001 | -7.52 | <0.001 | -1.02 | <0.001 | 8.00 | <0.001 | 1.73 | <0.001 |
| CingGy ventral area 23 L | Spos | 193.94 | <0.001 | 0.58 | <0.001 | -13.77 | <0.001 | -3.13 | <0.001 | -11.15 | <0.001 | -1.56 | <0.001 | 7.84 | <0.001 | 1.59 | <0.001 |
| CingGy ventral area 23 R | Spos | 223.40 | <0.001 | 0.61 | <0.001 | -15.28 | <0.001 | -3.45 | <0.001 | -10.40 | <0.001 | -1.46 | <0.001 | 9.29 | <0.001 | 1.85 | <0.001 |
| CingGy caudodorsal area 24 L | Spos | 217.16 | <0.001 | 0.61 | <0.001 | -14.29 | <0.001 | -3.23 | <0.001 | -7.96 | <0.001 | -1.08 | <0.001 | 10.38 | <0.001 | 2.17 | <0.001 |
| CingGy caudodorsal area 24 R | Spos | 215.77 | <0.001 | 0.61 | <0.001 | -14.97 | <0.001 | -3.34 | <0.001 | -8.16 | <0.001 | -1.13 | <0.001 | 10.33 | <0.001 | 2.09 | <0.001 |
| CingGy caudal area 23 L | Spos | 341.37 | <0.001 | 0.71 | <0.001 | -19.73 | <0.001 | -4.17 | <0.001 | -9.65 | <0.001 | -1.31 | <0.001 | 13.94 | <0.001 | 2.79 | <0.001 |
| CingGy caudal area 23 R | Spos | 296.83 | <0.001 | 0.68 | <0.001 | -17.54 | <0.001 | -3.85 | <0.001 | -9.58 | <0.001 | -1.31 | <0.001 | 12.28 | <0.001 | 2.50 | <0.001 |
| CingGy subgenual area 32 R | Spos | 108.94 | <0.001 | 0.44 | <0.001 | -9.94 | <0.001 | -2.31 | <0.001 | -7.51 | <0.001 | -1.04 | <0.001 | 6.33 | <0.001 | 1.32 | <0.001 |
| MedVentOC caudalingual gyrus L | Spos | 244.39 | <0.001 | 0.63 | <0.001 | -16.51 | <0.001 | -3.52 | <0.001 | -11.28 | <0.001 | -1.54 | <0.001 | 9.93 | <0.001 | 1.98 | <0.001 |
| MedVentOCostral cuneus gyrus L | Spos | 249.28 | <0.001 | 0.64 | <0.001 | -15.55 | <0.001 | -3.38 | <0.001 | -13.18 | <0.001 | -1.76 | <0.001 | 9.11 | <0.001 | 1.90 | <0.001 |
| MedVentOCostral cuneus gyrus R | Spos | 263.35 | <0.001 | 0.65 | <0.001 | -16.45 | <0.001 | -3.54 | <0.001 | -12.78 | <0.001 | -1.72 | <0.001 | 9.75 | <0.001 | 2.00 | <0.001 |
| MedVentOCostralingual gyrus L | Spos | 169.05 | <0.001 | 0.55 | <0.001 | -13.01 | <0.001 | -2.81 | <0.001 | -10.28 | <0.001 | -1.38 | <0.001 | 7.81 | <0.001 | 1.62 | <0.001 |
| MedVentOCostralingual gyrus R | Spos | 195.10 | <0.001 | 0.58 | <0.001 | -13.36 | <0.001 | -3.00 | <0.001 | -12.04 | <0.001 | -1.63 | <0.001 | 7.69 | <0.001 | 1.61 | <0.001 |
| MedVentOC ventromed parietocc sulcus L | Spos | 301.66 | <0.001 | 0.68 | <0.001 | -18.26 | <0.001 | -3.81 | <0.001 | -13.34 | <0.001 | -1.79 | <0.001 | 10.83 | <0.001 | 2.19 | <0.001 |
| MedVentOC ventromed parietocc sulcus R | Spos | 304.45 | <0.001 | 0.68 | <0.001 | -18.49 | <0.001 | -3.89 | <0.001 | -13.40 | <0.001 | -1.82 | <0.001 | 10.70 | <0.001 | 2.13 | <0.001 |
| Amyateral amygdala L | Spos | 96.60 | <0.001 | 0.41 | <0.001 | -9.96 | <0.001 | -2.17 | <0.001 | -7.94 | <0.001 | -1.08 | <0.001 | 5.67 | <0.001 | 1.15 | <0.001 |
| Amyateral amygdala R | Spos | 105.44 | <0.001 | 0.43 | <0.001 | -10.14 | <0.001 | -2.22 | <0.001 | -7.99 | <0.001 | -1.07 | <0.001 | 6.22 | <0.001 | 1.30 | <0.001 |
| Hippostral hipp L | Spos | 122.50 | <0.001 | 0.47 | <0.001 | -11.24 | <0.001 | -2.44 | <0.001 | -9.78 | <0.001 | -1.34 | <0.001 | 5.81 | <0.001 | 1.16 | <0.001 |
| Hipp caudal hipp L | Spos | 139.93 | <0.001 | 0.50 | <0.001 | -12.15 | <0.001 | -2.66 | <0.001 | -8.64 | <0.001 | -1.19 | <0.001 | 7.34 | <0.001 | 1.48 | <0.001 |
| Hipp caudal hipp R | Spos | 161.99 | <0.001 | 0.54 | <0.001 | -13.12 | <0.001 | -2.78 | <0.001 | -10.38 | <0.001 | -1.40 | <0.001 | 7.47 | <0.001 | 1.51 | <0.001 |
| BasGang ventral caudate L | Spos | 149.03 | <0.001 | 0.51 | <0.001 | -12.88 | <0.001 | -2.82 | <0.001 | -6.46 | <0.001 | -0.90 | <0.001 | 8.88 | <0.001 | 1.76 | <0.001 |
| BasGang ventral caudate R | Spos | 159.16 | <0.001 | 0.53 | <0.001 | -12.57 | <0.001 | -2.84 | <0.001 | -7.34 | <0.001 | -1.01 | <0.001 | 8.65 | <0.001 | 1.77 | <0.001 |
| BasGang globus pallidus L | Spos | 131.94 | <0.001 | 0.48 | <0.001 | -11.03 | <0.001 | -2.53 | <0.001 | -6.00 | <0.001 | -0.82 | <0.001 | 8.08 | <0.001 | 1.69 | <0.001 |
| BasGang globus pallidus R | Spos | 125.99 | <0.001 | 0.47 | <0.001 | -11.05 | <0.001 | -2.50 | <0.001 | -5.95 | <0.001 | -0.82 | <0.001 | 7.95 | <0.001 | 1.64 | <0.001 |
| BasGang nucleus accumbens L | Spos | 102.09 | <0.001 | 0.42 | <0.001 | -10.30 | <0.001 | -2.29 | <0.001 | -4.83 | <0.001 | -0.67 | <0.001 | 7.52 | <0.001 | 1.52 | <0.001 |
| BasGang ventromedial putamen L | Spos | 175.13 | <0.001 | 0.55 | <0.001 | -12.95 | <0.001 | -2.92 | <0.001 | -7.65 | <0.001 | -1.04 | <0.001 | 9.12 | <0.001 | 1.90 | <0.001 |
| BasGang ventromedial putamen R | Spos | 152.74 | <0.001 | 0.52 | <0.001 | -11.71 | <0.001 | -2.70 | <0.001 | -7.07 | <0.001 | -0.97 | <0.001 | 8.39 | <0.001 | 1.77 | <0.001 |
| BasGang dorsal caudate L | Spos | 202.04 | <0.001 | 0.59 | <0.001 | -14.72 | <0.001 | -3.26 | <0.001 | -8.11 | <0.001 | -1.13 | <0.001 | 9.96 | <0.001 | 1.99 | <0.001 |
| BasGang dorsal caudate R | Spos | 186.84 | <0.001 | 0.57 | <0.001 | -12.98 | <0.001 | -3.02 | <0.001 | -8.34 | <0.001 | -1.15 | <0.001 | 8.97 | <0.001 | 1.88 | <0.001 |
| BasGang dorsolat putamen L | Spos | 153.04 | <0.001 | 0.52 | <0.001 | -11.88 | <0.001 | -2.75 | <0.001 | -6.13 | <0.001 | -0.85 | <0.001 | 8.76 | <0.001 | 1.82 | <0.001 |
| BasGang dorsolat putamen R | Spos | 151.04 | <0.001 | 0.52 | <0.001 | -12.03 | <0.001 | -2.76 | <0.001 | -7.44 | <0.001 | -1.03 | <0.001 | 8.18 | <0.001 | 1.68 | <0.001 |
| Thal medial prefront thalamus L | Spos | 218.26 | <0.001 | 0.61 | <0.001 | -15.96 | <0.001 | -3.34 | <0.001 | -5.67 | <0.001 | -0.77 | <0.001 | 12.27 | <0.001 | 2.43 | <0.001 |
| Thal medial prefront thalamus R | Spos | 238.24 | <0.001 | 0.63 | <0.001 | -17.00 | <0.001 | -3.48 | <0.001 | -6.24 | <0.001 | -0.84 | <0.001 | 12.93 | <0.001 | 2.55 | <0.001 |
| Thal premotor thalamus L | Spos | 53.91 | <0.001 | 0.28 | <0.001 | -6.93 | <0.001 | -1.60 | <0.001 | -4.40 | <0.001 | -0.60 | <0.001 | 4.90 | <0.001 | 1.04 | <0.001 |
| Thal premotor thalamus R | Spos | 128.74 | <0.001 | 0.48 | <0.001 | -11.46 | <0.001 | -2.53 | <0.001 | -5.48 | <0.001 | -0.75 | <0.001 | 8.45 | <0.001 | 1.73 | <0.001 |
| Thal sensory thalamus L | Spos | 99.98 | <0.001 | 0.42 | <0.001 | -10.83 | <0.001 | -2.29 | <0.001 | -5.17 | <0.001 | -0.71 | <0.001 | 7.56 | <0.001 | 1.49 | <0.001 |
| Thal sensory thalamus R | Spos | 110.29 | <0.001 | 0.44 | <0.001 | -10.74 | <0.001 | -2.42 | <0.001 | -4.95 | <0.001 | -0.69 | <0.001 | 7.75 | <0.001 | 1.55 | <0.001 |
| Thalostral temp thalamus L | Spos | 177.86 | <0.001 | 0.56 | <0.001 | -12.69 | <0.001 | -2.97 | <0.001 | -6.97 | <0.001 | -0.97 | <0.001 | 9.23 | <0.001 | 1.93 | <0.001 |
| Thalostral temp thalamus R | Spos | 102.49 | <0.001 | 0.42 | <0.001 | -9.65 | <0.001 | -2.24 | <0.001 | -6.10 | <0.001 | -0.84 | <0.001 | 6.70 | <0.001 | 1.40 | <0.001 |
| Thal posterior parietal thalamus L | Spos | 192.20 | <0.001 | 0.58 | <0.001 | -15.29 | <0.001 | -3.25 | <0.001 | -6.02 | <0.001 | -0.83 | <0.001 | 11.07 | <0.001 | 2.14 | <0.001 |
| Thal posterior parietal thalamus R | Spos | 229.91 | <0.001 | 0.62 | <0.001 | -15.73 | <0.001 | -3.40 | <0.001 | -7.16 | <0.001 | -0.97 | <0.001 | 11.59 | <0.001 | 2.35 | <0.001 |
| Thal occipital thalamus L | Spos | 199.32 | <0.001 | 0.59 | <0.001 | -15.66 | <0.001 | -3.35 | <0.001 | -7.06 | <0.001 | -0.99 | <0.001 | 10.70 | <0.001 | 2.06 | <0.001 |
| Thal occipital thalamus R | Spos | 171.43 | <0.001 | 0.55 | <0.001 | -13.33 | <0.001 | -2.95 | <0.001 | -7.25 | <0.001 | -1.00 | <0.001 | 9.29 | <0.001 | 1.89 | <0.001 |
| Thal caudal temporal thalamus L | Spos | 164.32 | <0.001 | 0.54 | <0.001 | -14.14 | <0.001 | -2.99 | <0.001 | -5.96 | <0.001 | -0.82 | <0.001 | 10.06 | <0.001 | 1.95 | <0.001 |
| Thal caudal temporal thalamus R | Spos | 135.57 | <0.001 | 0.49 | <0.001 | -11.20 | <0.001 | -2.57 | <0.001 | -7.27 | <0.001 | -1.00 | <0.001 | 7.68 | <0.001 | 1.61 | <0.001 |
| Thalateral prefrontal thalamus L | Spos | 138.96 | <0.001 | 0.50 | <0.001 | -12.48 | <0.001 | -2.70 | <0.001 | -5.80 | <0.001 | -0.80 | <0.001 | 8.90 | <0.001 | 1.77 | <0.001 |
| Thalateral prefrontal thalamus R | Spos | 208.47 | <0.001 | 0.60 | <0.001 | -15.83 | <0.001 | -3.38 | <0.001 | -5.70 | <0.001 | -0.79 | <0.001 | 11.74 | <0.001 | 2.27 | <0.001 |
| Buckner cerebellar networks 1 | Spos | 140.83 | <0.001 | 0.50 | <0.001 | -11.19 | <0.001 | -2.64 | <0.001 | -9.78 | <0.001 | -1.38 | <0.001 | 6.45 | <0.001 | 1.33 | <0.001 |
| Buckner cerebellar networks 2 | Spos | 105.29 | <0.001 | 0.43 | <0.001 | -10.83 | <0.001 | -2.34 | <0.001 | -5.76 | <0.001 | -0.79 | <0.001 | 7.39 | <0.001 | 1.48 | <0.001 |
| Buckner cerebellar networks 4 | Spos | 199.60 | <0.001 | 0.59 | <0.001 | -15.87 | <0.001 | -3.30 | <0.001 | -7.60 | <0.001 | -1.05 | <0.001 | 10.67 | <0.001 | 2.05 | <0.001 |
| Buckner cerebellar networks 5 | Spos | 53.76 | <0.001 | 0.28 | <0.001 | -7.08 | <0.001 | -1.66 | <0.001 | -5.39 | <0.001 | -0.76 | <0.001 | 4.33 | <0.001 | 0.89 | <0.001 |
| Buckner cerebellar networks 9 | Spos | 164.42 | <0.001 | 0.54 | <0.001 | -13.50 | <0.001 | -2.90 | <0.001 | -5.77 | <0.001 | -0.79 | <0.001 | 9.99 | <0.001 | 2.00 | <0.001 |
| SFG medial frontal area 8 L | Sneg | 58.09 | <0.001 | 0.29 | <0.001 | 6.27 | <0.001 | 0.89 | <0.001 | -7.04 | <0.001 | -1.05 | <0.001 | -10.21 | <0.001 | -1.37 | <0.001 |
| SFG medial frontal area 8 R | Sneg | 45.68 | <0.001 | 0.25 | <0.001 | 8.36 | <0.001 | 1.04 | <0.001 | -5.61 | <0.001 | -0.83 | <0.001 | -10.03 | <0.001 | -1.31 | <0.001 |
| SFG dorsolateral area 6 L | Sneg | 47.36 | <0.001 | 0.25 | <0.001 | 6.86 | <0.001 | 0.92 | <0.001 | -5.97 | <0.001 | -0.88 | <0.001 | -9.97 | <0.001 | -1.35 | <0.001 |
| SFG medial area 6 L | Sneg | 51.53 | <0.001 | 0.27 | <0.001 | 6.88 | <0.001 | 0.92 | <0.001 | -6.41 | <0.001 | -0.95 | <0.001 | -10.15 | <0.001 | -1.35 | <0.001 |
| SFG medial area 6 R | Sneg | 57.35 | <0.001 | 0.29 | <0.001 | 8.51 | <0.001 | 1.06 | <0.001 | -6.63 | <0.001 | -0.99 | <0.001 | -10.72 | <0.001 | -1.39 | <0.001 |
| SFG medial area 9 L | Sneg | 51.07 | <0.001 | 0.27 | <0.001 | 7.44 | <0.001 | 0.97 | <0.001 | -5.99 | <0.001 | -0.85 | <0.001 | -11.15 | <0.001 | -1.53 | <0.001 |
| SFG medial area 9 R | Sneg | 41.82 | <0.001 | 0.23 | <0.001 | 7.34 | <0.001 | 1.01 | <0.001 | -4.93 | <0.001 | -0.70 | <0.001 | -9.98 | <0.001 | -1.41 | <0.001 |
| IFG dorsolateral area 44 R | Sneg | 33.23 | <0.001 | 0.19 | <0.001 | 8.62 | <0.001 | 1.07 | <0.001 | -4.16 | <0.001 | -0.61 | <0.001 | -8.96 | <0.001 | -1.18 | <0.001 |
| IFG opercular area 44 L | Sneg | 47.01 | <0.001 | 0.25 | <0.001 | 3.71 | <0.001 | 0.57 | <0.001 | -6.81 | <0.001 | -1.01 | <0.001 | -8.63 | <0.001 | -1.21 | <0.001 |
| IFG opercular area 44 R | Sneg | 51.76 | <0.001 | 0.27 | <0.001 | 4.50 | <0.001 | 0.70 | <0.001 | -6.96 | <0.001 | -1.03 | <0.001 | -9.26 | <0.001 | -1.29 | <0.001 |
| PreCG area 4 upperimb 4 L | Sneg | 28.86 | <0.001 | 0.17 | <0.001 | 6.40 | <0.001 | 0.82 | <0.001 | -4.12 | <0.001 | -0.58 | <0.001 | -8.76 | <0.001 | -1.21 | <0.001 |
| ParaCL area 1/2/3owerimb R | Sneg | 39.61 | <0.001 | 0.22 | <0.001 | 5.62 | <0.001 | 0.73 | <0.001 | -5.76 | <0.001 | -0.85 | <0.001 | -8.86 | <0.001 | -1.18 | <0.001 |
| ParaCL area 4owerimb L | Sneg | 34.27 | <0.001 | 0.20 | <0.001 | 7.37 | <0.001 | 0.87 | <0.001 | -4.71 | <0.001 | -0.68 | <0.001 | -9.40 | <0.001 | -1.24 | <0.001 |
| ParaCL area 4owerimb R | Sneg | 41.94 | <0.001 | 0.23 | <0.001 | 7.85 | <0.001 | 0.94 | <0.001 | -5.27 | <0.001 | -0.76 | <0.001 | -10.39 | <0.001 | -1.38 | <0.001 |
| FusGy medioventral area 37 L | Sneg | 21.90 | <0.001 | 0.13 | <0.001 | 5.52 | <0.001 | 0.65 | <0.001 | -3.65 | <0.001 | -0.49 | <0.001 | -8.72 | <0.001 | -1.20 | <0.001 |
| FusGy medioventral area 37 R | Sneg | 26.72 | <0.001 | 0.16 | <0.001 | 5.25 | <0.001 | 0.62 | <0.001 | -4.50 | <0.001 | -0.61 | <0.001 | -9.11 | <0.001 | -1.25 | <0.001 |
| ParHipGyateral post PH area L | Sneg | 75.70 | <0.001 | 0.35 | <0.001 | 2.62 | 0.01 | 0.41 | 0.01 | -9.20 | <0.001 | -1.37 | <0.001 | -10.16 | <0.001 | -1.42 | <0.001 |
| ParHipGyateral post PH area R | Sneg | 42.16 | <0.001 | 0.23 | <0.001 | 1.17 | 0.24 | 0.19 | 0.25 | -7.36 | <0.001 | -1.06 | <0.001 | -7.19 | <0.001 | -1.08 | <0.001 |
| ParHipGy Tharea medialPHG L | Sneg | 33.44 | <0.001 | 0.19 | <0.001 | -0.78 | 0.44 | -0.19 | 0.44 | -8.07 | <0.001 | -1.22 | <0.001 | -3.71 | <0.001 | -0.69 | <0.001 |
| ParHipGy Tharea medialPHG R | Sneg | 35.29 | <0.001 | 0.20 | <0.001 | -0.08 | 0.94 | -0.02 | 0.94 | -7.46 | <0.001 | -1.08 | <0.001 | -5.25 | <0.001 | -0.88 | <0.001 |
| PostSupTempSulc caudopost sup temp sulc L | Sneg | 27.36 | <0.001 | 0.16 | <0.001 | 5.20 | <0.001 | 0.69 | <0.001 | -4.45 | <0.001 | -0.63 | <0.001 | -8.11 | <0.001 | -1.13 | <0.001 |
| SPLateral area 5 L | Sneg | 21.64 | <0.001 | 0.13 | <0.001 | 4.15 | <0.001 | 0.70 | <0.001 | -3.65 | <0.001 | -0.52 | <0.001 | -6.38 | <0.001 | -1.01 | <0.001 |
| SPLateral area 5 R | Sneg | 26.70 | <0.001 | 0.16 | <0.001 | 7.30 | <0.001 | 0.88 | <0.001 | -3.36 | <0.001 | -0.46 | <0.001 | -9.11 | <0.001 | -1.25 | <0.001 |
| SPL intrapariet area 7 R | Sneg | 22.40 | <0.001 | 0.14 | <0.001 | 5.35 | <0.001 | 0.82 | <0.001 | -2.97 | <0.001 | -0.41 | <0.001 | -7.12 | <0.001 | -1.11 | <0.001 |
| PreCun medial area 7 L | Sneg | 47.55 | <0.001 | 0.25 | <0.001 | 8.55 | <0.001 | 0.96 | <0.001 | -5.76 | <0.001 | -0.83 | <0.001 | -11.10 | <0.001 | -1.44 | <0.001 |
| PreCun medial area 7 R | Sneg | 39.22 | <0.001 | 0.22 | <0.001 | 8.43 | <0.001 | 0.99 | <0.001 | -4.81 | <0.001 | -0.69 | <0.001 | -10.23 | <0.001 | -1.35 | <0.001 |
| PreCun medial area 5 L | Sneg | 38.76 | <0.001 | 0.22 | <0.001 | 7.86 | <0.001 | 0.86 | <0.001 | -5.41 | <0.001 | -0.80 | <0.001 | -9.28 | <0.001 | -1.18 | <0.001 |
| PreCun medial area 5 R | Sneg | 45.13 | <0.001 | 0.24 | <0.001 | 8.30 | <0.001 | 0.94 | <0.001 | -5.89 | <0.001 | -0.88 | <0.001 | -9.62 | <0.001 | -1.23 | <0.001 |
| PreCun dorsomed parietocc sulcus L | Sneg | 57.29 | <0.001 | 0.29 | <0.001 | 3.36 | <0.001 | 0.58 | <0.001 | -7.61 | <0.001 | -1.15 | <0.001 | -8.83 | <0.001 | -1.24 | <0.001 |
| PreCun dorsomed parietocc sulcus R | Sneg | 74.02 | <0.001 | 0.35 | <0.001 | 4.01 | <0.001 | 0.64 | <0.001 | -8.64 | <0.001 | -1.30 | <0.001 | -10.33 | <0.001 | -1.42 | <0.001 |
| PreCun area 31 L | Sneg | 54.01 | <0.001 | 0.28 | <0.001 | 3.44 | <0.001 | 0.55 | <0.001 | -7.50 | <0.001 | -1.10 | <0.001 | -9.07 | <0.001 | -1.31 | <0.001 |
| PreCun area 31 R | Sneg | 64.00 | <0.001 | 0.31 | <0.001 | 4.73 | <0.001 | 0.68 | <0.001 | -7.91 | <0.001 | -1.16 | <0.001 | -10.56 | <0.001 | -1.46 | <0.001 |
| InsL hypergranular insular L | Sneg | 41.60 | <0.001 | 0.23 | <0.001 | 1.48 | 0.14 | 0.27 | 0.15 | -7.12 | <0.001 | -1.05 | <0.001 | -6.86 | <0.001 | -1.05 | <0.001 |
| InsL hypergranular insular R | Sneg | 44.43 | <0.001 | 0.24 | <0.001 | 1.86 | 0.07 | 0.33 | 0.07 | -7.10 | <0.001 | -1.06 | <0.001 | -7.37 | <0.001 | -1.07 | <0.001 |
| InsL ventral agranular insular L | Sneg | 29.21 | <0.001 | 0.17 | <0.001 | 0.03 | 0.97 | 0.01 | 0.97 | -6.62 | <0.001 | -0.98 | <0.001 | -4.74 | <0.001 | -0.78 | <0.001 |
| InsL ventral agranular insular R | Sneg | 23.43 | <0.001 | 0.14 | <0.001 | -1.74 | 0.09 | -0.42 | 0.09 | -7.15 | <0.001 | -1.06 | <0.001 | -2.18 | 0.03 | -0.41 | 0.03 |
| InsL dorsolat agranular insular L | Sneg | 50.02 | <0.001 | 0.26 | <0.001 | 2.90 | <0.001 | 0.49 | 0.01 | -7.33 | <0.001 | -1.08 | <0.001 | -8.39 | <0.001 | -1.23 | <0.001 |
| InsL dorsolat agranular insular R | Sneg | 54.77 | <0.001 | 0.28 | <0.001 | 4.09 | <0.001 | 0.61 | <0.001 | -7.38 | <0.001 | -1.09 | <0.001 | -9.55 | <0.001 | -1.33 | <0.001 |
| InsL ventral granular insular L | Sneg | 43.07 | <0.001 | 0.23 | <0.001 | 0.54 | 0.59 | 0.12 | 0.59 | -7.51 | <0.001 | -1.14 | <0.001 | -6.07 | <0.001 | -0.95 | <0.001 |
| InsL ventral granular insular R | Sneg | 46.90 | <0.001 | 0.25 | <0.001 | 2.44 | 0.02 | 0.40 | 0.02 | -7.14 | <0.001 | -1.06 | <0.001 | -8.01 | <0.001 | -1.13 | <0.001 |
| InsL dorsolat granular insular L | Sneg | 32.09 | <0.001 | 0.19 | <0.001 | 3.14 | <0.001 | 0.46 | <0.001 | -5.84 | <0.001 | -0.81 | <0.001 | -7.93 | <0.001 | -1.20 | <0.001 |
| InsL dorsolat granular insular R | Sneg | 36.53 | <0.001 | 0.21 | <0.001 | 1.46 | 0.15 | 0.29 | 0.15 | -7.13 | <0.001 | -1.02 | <0.001 | -6.19 | <0.001 | -1.06 | <0.001 |
| InsL dorsal dysgranular insular L | Sneg | 44.03 | <0.001 | 0.24 | <0.001 | 2.26 | 0.03 | 0.38 | 0.03 | -7.00 | <0.001 | -1.04 | <0.001 | -7.66 | <0.001 | -1.11 | <0.001 |
| InsL dorsal dysgranular insular R | Sneg | 50.10 | <0.001 | 0.26 | <0.001 | 3.64 | <0.001 | 0.60 | <0.001 | -7.06 | <0.001 | -1.05 | <0.001 | -8.71 | <0.001 | -1.24 | <0.001 |
| CingGy dorsal area 23 L | Sneg | 58.96 | <0.001 | 0.30 | <0.001 | 5.97 | <0.001 | 0.80 | <0.001 | -7.23 | <0.001 | -1.06 | <0.001 | -10.67 | <0.001 | -1.44 | <0.001 |
| CingGy dorsal area 23 R | Sneg | 56.59 | <0.001 | 0.29 | <0.001 | 5.21 | <0.001 | 0.71 | <0.001 | -7.25 | <0.001 | -1.07 | <0.001 | -10.12 | <0.001 | -1.36 | <0.001 |
| CingGyostroventral area 24 L | Sneg | 30.93 | <0.001 | 0.18 | <0.001 | 2.18 | 0.03 | 0.35 | 0.03 | -5.76 | <0.001 | -0.86 | <0.001 | -6.64 | <0.001 | -0.94 | <0.001 |
| CingGy pregenual area 32 L | Sneg | 74.36 | <0.001 | 0.35 | <0.001 | 2.42 | 0.02 | 0.43 | 0.02 | -9.05 | <0.001 | -1.37 | <0.001 | -9.48 | <0.001 | -1.35 | <0.001 |
| CingGy pregenual area 32 R | Sneg | 75.07 | <0.001 | 0.35 | <0.001 | 2.95 | <0.001 | 0.48 | <0.001 | -8.84 | <0.001 | -1.36 | <0.001 | -9.78 | <0.001 | -1.32 | <0.001 |
| CingGy ventral area 23 L | Sneg | 69.08 | <0.001 | 0.33 | <0.001 | 3.73 | <0.001 | 0.58 | <0.001 | -8.40 | <0.001 | -1.27 | <0.001 | -9.99 | <0.001 | -1.36 | <0.001 |
| CingGy ventral area 23 R | Sneg | 75.36 | <0.001 | 0.35 | <0.001 | 4.17 | <0.001 | 0.63 | <0.001 | -8.71 | <0.001 | -1.31 | <0.001 | -10.55 | <0.001 | -1.42 | <0.001 |
| CingGy caudodorsal area 24 L | Sneg | 67.93 | <0.001 | 0.33 | <0.001 | 5.64 | <0.001 | 0.75 | <0.001 | -7.98 | <0.001 | -1.21 | <0.001 | -10.53 | <0.001 | -1.38 | <0.001 |
| CingGy caudodorsal area 24 R | Sneg | 59.79 | <0.001 | 0.30 | <0.001 | 5.62 | <0.001 | 0.72 | <0.001 | -7.46 | <0.001 | -1.11 | <0.001 | -10.43 | <0.001 | -1.37 | <0.001 |
| CingGy caudal area 23 L | Sneg | 37.31 | <0.001 | 0.21 | <0.001 | 7.87 | <0.001 | 0.93 | <0.001 | -4.81 | <0.001 | -0.69 | <0.001 | -9.97 | <0.001 | -1.32 | <0.001 |
| CingGy caudal area 23 R | Sneg | 40.41 | <0.001 | 0.22 | <0.001 | 5.94 | <0.001 | 0.79 | <0.001 | -5.71 | <0.001 | -0.85 | <0.001 | -8.97 | <0.001 | -1.20 | <0.001 |
| CingGy subgenual area 32 R | Sneg | 36.18 | <0.001 | 0.20 | <0.001 | -0.27 | 0.79 | -0.06 | 0.79 | -7.86 | <0.001 | -1.16 | <0.001 | -4.62 | <0.001 | -0.82 | <0.001 |
| MedVentOC caudalingual gyrus L | Sneg | 25.24 | <0.001 | 0.15 | <0.001 | 6.19 | <0.001 | 0.72 | <0.001 | -3.98 | <0.001 | -0.55 | <0.001 | -8.73 | <0.001 | -1.16 | <0.001 |
| MedVentOCostral cuneus gyrus L | Sneg | 26.44 | <0.001 | 0.16 | <0.001 | 3.81 | <0.001 | 0.55 | <0.001 | -4.83 | <0.001 | -0.69 | <0.001 | -7.31 | <0.001 | -1.05 | <0.001 |
| MedVentOCostral cuneus gyrus R | Sneg | 29.95 | <0.001 | 0.18 | <0.001 | 3.90 | <0.001 | 0.55 | <0.001 | -5.24 | <0.001 | -0.74 | <0.001 | -7.89 | <0.001 | -1.13 | <0.001 |
| MedVentOCostralingual gyrus L | Sneg | 16.02 | <0.001 | 0.10 | <0.001 | 2.42 | 0.02 | 0.37 | 0.02 | -4.04 | <0.001 | -0.55 | <0.001 | -5.63 | <0.001 | -0.89 | <0.001 |
| MedVentOCostralingual gyrus R | Sneg | 20.20 | <0.001 | 0.13 | <0.001 | 3.64 | <0.001 | 0.48 | <0.001 | -4.20 | <0.001 | -0.56 | <0.001 | -7.41 | <0.001 | -1.09 | <0.001 |
| MedVentOC ventromed parietocc sulcus L | Sneg | 37.94 | <0.001 | 0.21 | <0.001 | 5.05 | <0.001 | 0.63 | <0.001 | -5.80 | <0.001 | -0.82 | <0.001 | -9.37 | <0.001 | -1.27 | <0.001 |
| MedVentOC ventromed parietocc sulcus R | Sneg | 44.51 | <0.001 | 0.24 | <0.001 | 4.57 | <0.001 | 0.61 | <0.001 | -6.51 | <0.001 | -0.93 | <0.001 | -9.56 | <0.001 | -1.32 | <0.001 |
| Amyateral amygdala L | Sneg | 35.21 | <0.001 | 0.20 | <0.001 | 1.80 | 0.08 | 0.31 | 0.08 | -6.46 | <0.001 | -0.94 | <0.001 | -6.72 | <0.001 | -1.02 | <0.001 |
| Amyateral amygdala R | Sneg | 26.32 | <0.001 | 0.16 | <0.001 | 1.19 | 0.24 | 0.22 | 0.24 | -5.84 | <0.001 | -0.85 | <0.001 | -5.44 | <0.001 | -0.87 | <0.001 |
| Hippostral hipp L | Sneg | 29.65 | <0.001 | 0.17 | <0.001 | 1.75 | 0.08 | 0.31 | 0.09 | -5.73 | <0.001 | -0.86 | <0.001 | -6.13 | <0.001 | -0.89 | <0.001 |
| Hipp caudal hipp L | Sneg | 55.48 | <0.001 | 0.28 | <0.001 | 1.35 | 0.18 | 0.23 | 0.18 | -8.04 | <0.001 | -1.21 | <0.001 | -7.99 | <0.001 | -1.13 | <0.001 |
| Hipp caudal hipp R | Sneg | 52.89 | <0.001 | 0.27 | <0.001 | 1.36 | 0.18 | 0.24 | 0.18 | -7.74 | <0.001 | -1.18 | <0.001 | -7.73 | <0.001 | -1.08 | <0.001 |
| BasGang ventral caudate L | Sneg | 60.34 | <0.001 | 0.30 | <0.001 | 2.31 | 0.02 | 0.36 | 0.02 | -8.21 | <0.001 | -1.22 | <0.001 | -9.09 | <0.001 | -1.26 | <0.001 |
| BasGang ventral caudate R | Sneg | 62.36 | <0.001 | 0.31 | <0.001 | 3.25 | <0.001 | 0.50 | <0.001 | -8.06 | <0.001 | -1.21 | <0.001 | -9.43 | <0.001 | -1.29 | <0.001 |
| BasGang globus pallidus L | Sneg | 42.66 | <0.001 | 0.23 | <0.001 | 1.66 | 0.10 | 0.32 | 0.10 | -7.07 | <0.001 | -1.06 | <0.001 | -6.91 | <0.001 | -1.04 | <0.001 |
| BasGang globus pallidus R | Sneg | 51.70 | <0.001 | 0.27 | <0.001 | 2.72 | 0.01 | 0.46 | 0.01 | -7.47 | <0.001 | -1.11 | <0.001 | -8.39 | <0.001 | -1.21 | <0.001 |
| BasGang nucleus accumbens L | Sneg | 60.98 | <0.001 | 0.30 | <0.001 | 0.85 | 0.40 | 0.14 | 0.40 | -8.47 | <0.001 | -1.28 | <0.001 | -8.38 | <0.001 | -1.15 | <0.001 |
| BasGang ventromedial putamen L | Sneg | 44.04 | <0.001 | 0.24 | <0.001 | 2.47 | 0.02 | 0.46 | 0.02 | -6.99 | <0.001 | -1.03 | <0.001 | -7.47 | <0.001 | -1.14 | <0.001 |
| BasGang ventromedial putamen R | Sneg | 52.25 | <0.001 | 0.27 | <0.001 | 3.69 | <0.001 | 0.55 | <0.001 | -7.31 | <0.001 | -1.07 | <0.001 | -9.26 | <0.001 | -1.30 | <0.001 |
| BasGang dorsal caudate L | Sneg | 41.75 | <0.001 | 0.23 | <0.001 | 3.84 | <0.001 | 0.61 | <0.001 | -6.30 | <0.001 | -0.94 | <0.001 | -8.21 | <0.001 | -1.15 | <0.001 |
| BasGang dorsal caudate R | Sneg | 42.43 | <0.001 | 0.23 | <0.001 | 2.77 | 0.01 | 0.45 | 0.01 | -6.69 | <0.001 | -0.99 | <0.001 | -7.83 | <0.001 | -1.11 | <0.001 |
| BasGang dorsolat putamen L | Sneg | 40.54 | <0.001 | 0.22 | <0.001 | 1.91 | 0.06 | 0.35 | 0.06 | -6.74 | <0.001 | -1.01 | <0.001 | -7.00 | <0.001 | -1.03 | <0.001 |
| BasGang dorsolat putamen R | Sneg | 25.65 | <0.001 | 0.15 | <0.001 | 1.31 | 0.20 | 0.26 | 0.20 | -5.92 | <0.001 | -0.85 | <0.001 | -5.14 | <0.001 | -0.88 | <0.001 |
| Thal medial prefront thalamus L | Sneg | 44.22 | <0.001 | 0.24 | <0.001 | 5.02 | <0.001 | 0.65 | <0.001 | -6.37 | <0.001 | -0.97 | <0.001 | -8.59 | <0.001 | -1.12 | <0.001 |
| Thal medial prefront thalamus R | Sneg | 44.78 | <0.001 | 0.24 | <0.001 | 7.63 | <0.001 | 0.83 | <0.001 | -6.05 | <0.001 | -0.91 | <0.001 | -9.52 | <0.001 | -1.21 | <0.001 |
| Thal premotor thalamus L | Sneg | 31.05 | <0.001 | 0.18 | <0.001 | 0.26 | 0.79 | 0.05 | 0.80 | -6.27 | <0.001 | -0.95 | <0.001 | -5.39 | <0.001 | -0.80 | <0.001 |
| Thal premotor thalamus R | Sneg | 51.44 | <0.001 | 0.27 | <0.001 | 5.35 | <0.001 | 0.73 | <0.001 | -6.83 | <0.001 | -1.03 | <0.001 | -9.45 | <0.001 | -1.25 | <0.001 |
| Thal sensory thalamus L | Sneg | 36.93 | <0.001 | 0.21 | <0.001 | 3.14 | <0.001 | 0.49 | <0.001 | -6.03 | <0.001 | -0.91 | <0.001 | -7.39 | <0.001 | -1.00 | <0.001 |
| Thal sensory thalamus R | Sneg | 62.16 | <0.001 | 0.31 | <0.001 | 1.67 | 0.10 | 0.33 | 0.10 | -8.47 | <0.001 | -1.29 | <0.001 | -8.07 | <0.001 | -1.20 | <0.001 |
| Thalostral temp thalamus L | Sneg | 55.30 | <0.001 | 0.28 | <0.001 | 5.68 | <0.001 | 0.70 | <0.001 | -7.15 | <0.001 | -1.05 | <0.001 | -10.34 | <0.001 | -1.36 | <0.001 |
| Thalostral temp thalamus R | Sneg | 47.47 | <0.001 | 0.25 | <0.001 | 0.04 | 0.97 | 0.01 | 0.97 | -7.88 | <0.001 | -1.19 | <0.001 | -6.51 | <0.001 | -0.97 | <0.001 |
| Thal posterior parietal thalamus L | Sneg | 33.32 | <0.001 | 0.19 | <0.001 | 3.92 | <0.001 | 0.61 | <0.001 | -5.48 | <0.001 | -0.83 | <0.001 | -7.30 | <0.001 | -0.99 | <0.001 |
| Thal posterior parietal thalamus R | Sneg | 45.89 | <0.001 | 0.25 | <0.001 | 2.57 | 0.01 | 0.46 | 0.01 | -6.97 | <0.001 | -1.05 | <0.001 | -7.71 | <0.001 | -1.11 | <0.001 |
| Thal occipital thalamus L | Sneg | 50.55 | <0.001 | 0.26 | <0.001 | 6.73 | <0.001 | 0.84 | <0.001 | -6.49 | <0.001 | -0.96 | <0.001 | -10.07 | <0.001 | -1.32 | <0.001 |
| Thal occipital thalamus R | Sneg | 55.03 | <0.001 | 0.28 | <0.001 | 3.80 | <0.001 | 0.57 | <0.001 | -7.39 | <0.001 | -1.12 | <0.001 | -9.10 | <0.001 | -1.23 | <0.001 |
| Thal caudal temporal thalamus L | Sneg | 42.40 | <0.001 | 0.23 | <0.001 | 4.31 | <0.001 | 0.57 | <0.001 | -6.34 | <0.001 | -0.95 | <0.001 | -8.50 | <0.001 | -1.12 | <0.001 |
| Thal caudal temporal thalamus R | Sneg | 43.18 | <0.001 | 0.24 | <0.001 | 1.63 | 0.11 | 0.28 | 0.11 | -7.11 | <0.001 | -1.06 | <0.001 | -7.29 | <0.001 | -1.06 | <0.001 |
| Thalateral prefrontal thalamus L | Sneg | 32.08 | <0.001 | 0.19 | <0.001 | 3.51 | <0.001 | 0.53 | <0.001 | -5.50 | <0.001 | -0.85 | <0.001 | -6.81 | <0.001 | -0.89 | <0.001 |
| Thalateral prefrontal thalamus R | Sneg | 47.56 | <0.001 | 0.25 | <0.001 | 5.51 | <0.001 | 0.73 | <0.001 | -6.53 | <0.001 | -0.99 | <0.001 | -8.97 | <0.001 | -1.17 | <0.001 |
| Buckner cerebellar networks 1 | Sneg | 20.79 | <0.001 | 0.13 | <0.001 | 2.31 | 0.02 | 0.42 | 0.02 | -4.67 | <0.001 | -0.66 | <0.001 | -5.49 | <0.001 | -0.90 | <0.001 |
| Buckner cerebellar networks 2 | Sneg | 45.98 | <0.001 | 0.25 | <0.001 | 2.37 | 0.02 | 0.37 | 0.02 | -7.07 | <0.001 | -1.06 | <0.001 | -7.98 | <0.001 | -1.11 | <0.001 |
| Buckner cerebellar networks 4 | Sneg | 37.26 | <0.001 | 0.21 | <0.001 | 3.84 | <0.001 | 0.52 | <0.001 | -5.98 | <0.001 | -0.90 | <0.001 | -7.88 | <0.001 | -1.05 | <0.001 |
| Buckner cerebellar networks 5 | Sneg | 23.86 | <0.001 | 0.15 | <0.001 | -0.91 | 0.37 | -0.19 | 0.37 | -6.72 | <0.001 | -0.97 | <0.001 | -3.38 | <0.001 | -0.60 | <0.001 |
| Buckner cerebellar networks 9 | Sneg | 34.86 | <0.001 | 0.20 | <0.001 | 1.92 | 0.06 | 0.39 | 0.06 | -6.42 | <0.001 | -0.95 | <0.001 | -6.20 | <0.001 | -1.01 | <0.001 |

For each Brainnetome label the corresponding region of interest (ROI) is reported Abbreviations: HC, healthy controls; SFG, superior frontal gyrus; MFG, medial frontal gyrus; IFG; inferior frontal gyrus; OrG, orbital gyrus; PreCG, precentral gyrus; ParaCL, paracentral lobule; STG, superior temporal gyrus; MTG, medial temporal gyrus; ITG, inferior temporal gyrus; FusGy, fusiform gyrus; ParHipGy, hippocampal part of the cingulate gyrus; PostSupTempSulc, posterior and superior temporal sulcus; Sneg, negative functional connectivity strength; SPL, superior parietal lobule; Spos, positive functional connectivity strength; IPL, inferior parietal lobule; PreCun, precuneus; PostCGy, posterior cingulate gyrus; Ins, insula; CingCy, cingulate gyrus; MedVenOC, medial ventral occipital cortex; LatOC, lateral occipital cortex; Amy, amygdala; Hipp, hippocampus; BasGang, basal ganglia; Thal, thalamus; L, left; R, right.
